# Essential Indicators of Quality in Primary Care Settings: An Evidence-Based, Structured, Expert Approach

**DOI:** 10.1101/2021.09.13.21262970

**Authors:** S.J. Hysong, K. Arredondo, A.M. Hughes, H.F. Lester, F.L. Oswald, L.A. Petersen, L. Woodard, E. Post, S. DePeralta, D.R. Murphy, J. McKnight, K. Nelson, P. Haidet

## Abstract

**Introduction:** Primary health care has a central role in the workings of the health care system and health of the American public. Thus, a high-performing, high-quality primary care system is essential. As a result, measurement frameworks are needed to assess the quality of the infrastructure, workforce configurations, and processes available in primary care practices due to the complexity of primary care. As part of a larger project supported by AHRQ (grant no. 1 R01 HS 025982), our research team reports the use of an evidence-based approach to compile a targeted set of existing care measures. These measures are prioritized according to their overall contribution and value to primary care. Within this paper, we describe the process by which the performance measures were selected and present the final set of measures resulting from the process.

**Defining Primary care:** The study centers around general primary care settings, which have been defined as, “the provision of integrated, accessible health care services by clinicians who are accountable for addressing a large majority of personal health care needs, developing a sustained partnership with patients, and practicing in the context of family and community.” (1)

**Using PROMES:** We adapted an evidence-based approach for measure development, The Productivity Measurement and Enhancement System, or ProMES, to select (modify, when appropriate) and rank existing primary care measures according to value to the primary care clinic. ProMES is a comprehensive performance measure development approach firmly grounded in motivational theory and performance measurement.(2, 3) Through a facilitated focus-group based process, these measures are defined, weighted, and prioritized to create indicators of both overall effectiveness and specific aspects of daily work. This alignment helps individuals and teams to focus their effort more clearly on the most important aspects of their work (i.e., clinical performance) resulting in greater productivity, reduced stress, and less waste of effort.(2, 3) We utilize the ProMES definition of *productivity*, which is how effectively an organization uses its resources to achieve its goals.

One unique feature of ProMES is the resulting measures include contingency curves, or non-linear functions that explicitly tie performance levels on a given measure to its contribution to the organization’s values; in this way, the application of ProMES yields a more nuanced approach to prioritizing work than simple linear weights, while allowing direct comparison(s) between measure(s).

**Results:** The design team identified three fundamental objectives for delivery of high-quality primary care. The design team also selected sixteen performance indicators from the 44 pre-vetted measures that already exist in three different data sources for primary care. One indicator, Team 2 Day Post Discharge Contact Ratio, was selected as an indicator for both Objective 2 and 3. In addition, contingency curves were created for each of the indicators using the contingency functions developed by the design team.

- *Objective 1*. Ensure patient has appropriate access to preventive, acute, or chronic health care services when needed.
  – Indicators:
    - New Primary Care Patient Average Wait Time in Days
    - Established Primary Care Patient Average Wait Time in Days
    - Average 3rd Next Available Appointment in PC Clinics
    - Total Inbound PC Secure Messages to Total Outbound PC Secure Messages (Ratio)
    - Urgent Care Utilization Rate
- *Objective 2.* Build a trusting, effective, sustained partnership between the health-care team, the patient, and his/her caregiver(s) towards shared goals.
  – Indicators:
    - Patient’s Satisfaction Rating of Primary Care Provider
    - Team 2 Day Post Discharge Contact Ratio
    - Patient-Centered Medical Home Stress Discussed
- *Objective 3.* Deliver safe and effective care that comprehensively addresses a given patient’s particular ecological, biological, and/or psychosocial needs.
  – Indicators:
    - Ambulatory Care Sensitive Conditions (ACSC) Hospitalizations Rate Per 1000 Patients
    - Diabetes Patients with HbA1c Poor Control
    - Diabetes Electronic Composite Measure
    - Statin Medication for Patients with Cardiovascular Disease
    - Controlling High Blood Pressure
    - Renal Testing for Nephropathy
    - Effective Continuation Phase Treatment for depression
    - Hospital-wide all cause 30-day Readmission Rate
    - Team 2 Day Post Discharge Contact Ratio

**Summary:** Performance measures selected as part of our modified-ProMES process assist in the implementation of targeted care quality measures prioritized in accordance to their value in primary care. By deriving high-value metrics, organized by care objective with numerically assigned prioritization, we anticipate the results of this paper will apply to a diverse set of stakeholders, including but not limited to policy-makers, primary care clinicians, and administrators in healthcare organizations. Our design team of nationally recognized SMEs joined together in a national panel that consists of diverse stakeholder groups to collectively identify three primary care team objectives, 16 indicators of primary care quality, and 13 indicators which require modification and further work to address gaps which exist in the primary care performance measurement domain. Measures selected as part of this study aim were constructed independent of clinic size or configuration, so that clinics of many configurations (e.g., public vs. private, large vs. small, rural vs urban, team-based vs. traditional) could benefit from their use.

Our measure set provides an actionable catalogue of measures that can serve as a first step toward interoperability of electronic health record systems. Future work toward this goal should address both logistical considerations (e.g., data capture, common data/programming language) and lingering measurement challenges, such as the best way to operationalize these measures for teams working in complex and shifting situations (e.g., rotating team members).

**Acronyms and Definitions:** 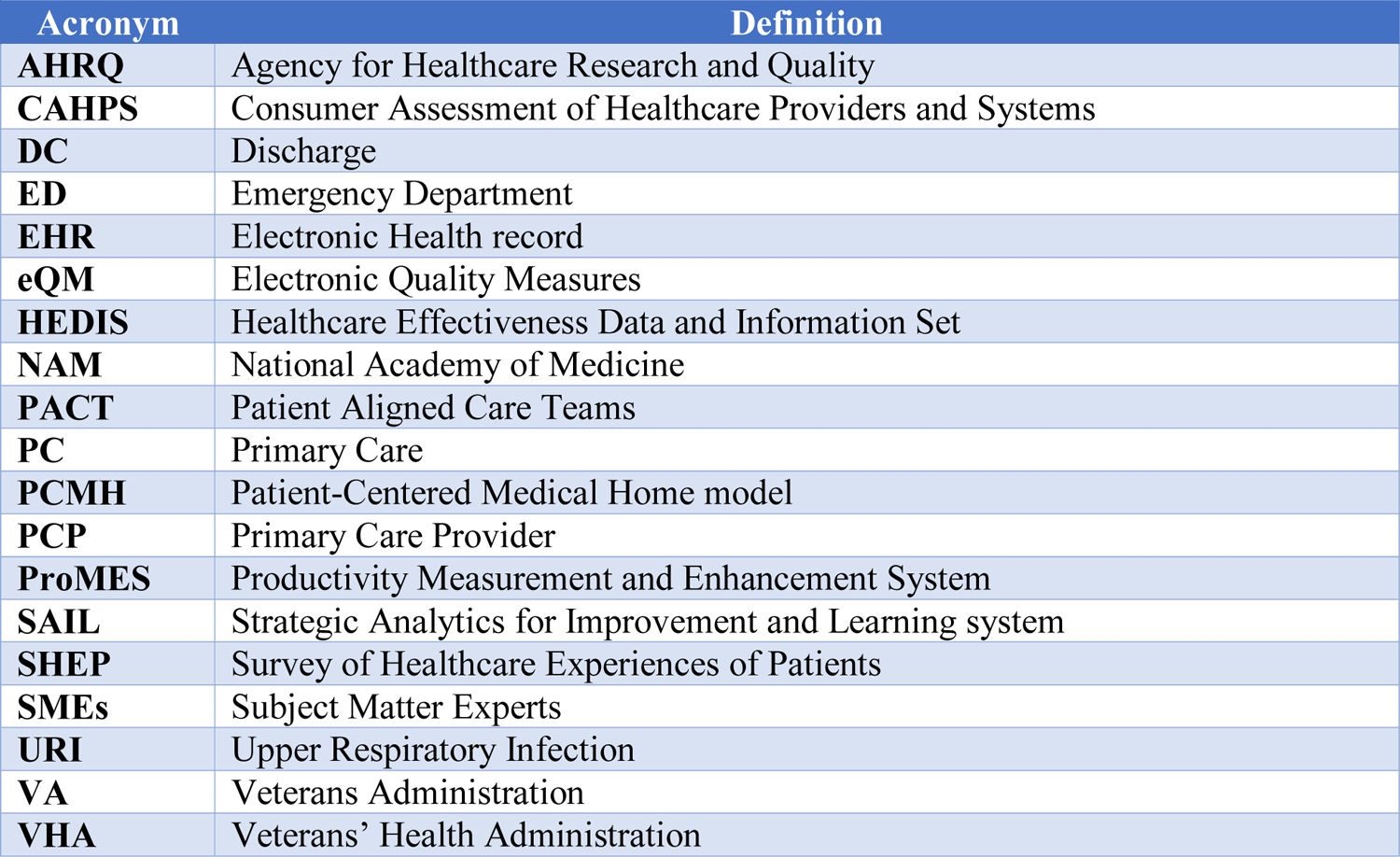

## Introduction

Primary health care provides the principal point of entry for patients into the American health care system, and is often the gateway for initially accessing most other specialties. Primary care involves the provision of integrated, accessible health care services to address a majority of common health care needs and maintain life-long partnerships with patients.(1) Because of its central role in the workings of the health care system and health of the American public, a high-performing, high-quality primary care system is essential. Thus, private and public entities alike have devoted considerable resources toward that goal.

Primary care delivery has continued to evolve over the last few decades facilitated by significant advances in medicine, technology, and policy. As a result, measurement frameworks are needed to assess the quality of the infrastructure, workforce configurations, and processes available in primary care practices due to the complexity of primary care. A plethora of measures currently exists to help monitor primary care activities, including the number of appointments, patient load, and clinical performance assessments of patient-related data (e.g., the Healthcare Effectiveness Data and Information Set (HEDIS)). Furthermore, new workforce configurations such as Patient Aligned Care Teams (PACT) -- the Veterans Health Administration’s (VHA) Patient-Centered Medical Home (PCMH) model -- have accelerated the development of measures to assess the quality of primary care (please see the table at the beginning of the document for a complete list of acronyms and definitions used in this document).

Despite advances in primary care delivery (e.g., delivered by teams, greater use of electronic health records and patient portals), the fundamental objectives of primary care (e.g., improving population health) and the nature of the work typically performed in primary care (e.g., patient assessment and treatment of common and chronic conditions, medication management, preventive care, coordination of care for complex patients) have generally remained unchanged.(4) As a result, this raises questions regarding the necessity of recent proliferation of novel quality measures and clinical performance measures. Given the exponential growth of readily available digital health information in the last decade, distinguishing meaningful, appropriate, and high-quality performance measures from irrelevant, unhelpful, or unrepresentative performance data quickly becomes a necessary yet time-demanding undertaking.

The proliferation of performance measures, of both high and low quality, bears unintended consequences on the primary care workforce; for example, in 2014, Kansagara and colleagues reported that primary care staff perceived that some performance measures: “1) led to delivery changes that were not always aligned with PACT principles, 2) did not accurately reflect patient-priorities, 3) represented an opportunity cost, 4) were imposed with little communication or transparency, and 5) were not well-adapted to team-based care.”(5) Powell and colleagues(6) further suggested that implementing additional performance measures generates non-value-added clinical burden, such as redundant documentation and a tendency to focus on actions that quickly improve scores, as opposed to actually improving patient outcomes. These actions, in turn, often result in inappropriate patient care and decreased patient-centeredness (e.g., lack of focus on patient concerns). Further complicating the generation of new measures, implementation of new quality care measures can disrupt workflow, adversely impacting dynamics within the primary care team. Prior work illustrates the vast range of unintended consequences for collecting new care quality and performance measures, ranging from redundant or unnecessary work to clinician dissatisfaction and burnout, leading to an exodus from primary care and worsening primary care provider shortages.

Such concerns suggest that assessing whether primary care practices are delivering on the fundamental promise and objectives of primary care as a domain, should be accomplished through a tailored set of measurable, accountable, prioritized primary care performance measures. As part of a larger project supported by AHRQ (grant no. 1 R01 HS 025982), our research team reports the use of an evidence-based approach to compile a targeted set of existing care measures. These measures are prioritized according to their overall contribution and value to primary care. Within this paper, we describe the process by which the performance measures were selected and present the final set of measures resulting from the process.

## Approach

### Overview

Ideally, scientific evidence informs the development, validation, and ultimately the application(s) of performance measures. Specifically, good performance measures propose criteria by which the performer is to be evaluated. A set of performance measures, however, can serve multiple purposes, including assessment of similar criteria across different settings (e.g., multiple organizational types) and different people (e.g., multiple subjects). Further, performance measures are adaptable in that they can serve more than one purpose or interpretation of their criteria. For example, blood pressure and heart rate measures (e.g., heart rate variability, beat-to-beat intervals) can indicate an individuals’ cardiovascular health.(7) However, this same set of measures in a healthy sample of adults can also be used to infer an individuals’ level of stress and experienced workload.(8, 9) Rather than flood the literature with additional measures – which are resource intensive to develop, validate, and integrate into clinical practice – we detail the adaptation of an evidence-based approach for measure development, (The Productivity Measurement and Enhancement System, or ProMES, described in more detail below), to select (modify, when appropriate) and rank existing primary care measures according to value to the primary care clinic. ProMES is a structured, comprehensive productivity improvement approach firmly grounded in motivational theory, the science of collective sensemaking, and performance measurement,(2,10,11) and has been used successfully in primary healthcare settings.(12) In utilizing this approach, we can more readily apply an evidence-based practice to guide selection and focus of primary care teams to impactful quality improvement efforts. To meet the objectives of this project, we convened a panel of nationally recognized Subject Matter Experts (SMEs, described in more detail below) in primary care and performance assessment. We employed ProMES to guide the SMEs through the process of identifying key objectives in the delivery of primary care, selecting performance measures from existing data sources indicative of whether the objectives in question were being achieved, and prioritizing the measure set by evaluating the value to primary care of maximizing performance in each indicator.

### Defining Primary Care

The study centers around general primary care settings, which have been defined as, “the provision of integrated, accessible health care services by clinicians who are accountable for addressing a large majority of personal health care needs, developing a sustained partnership with patients, and practicing in the context of family and community.”(1) Since this classic definition was published by the National Academy of Medicine (NAM), others have expanded on the concept. Despite expansions such as empanelment(13, 14) and data-driven improvement,(15) a recent AHRQ white paper on redefining primary care shows increasing consensus on the components of high-quality primary care. All major frameworks cited by this report agree that high-quality primary care should (a) deliver prompt access to care; (b) be delivered by a team; (c) engage patients as partners in the team; (d) carry out population management; and (e) deliver comprehensive, coordinated care, all elements that are present in the original NAM definition. Thus, the methods and procedures described herein rely on the classic NAM definition as its foundation.

### Using ProMES to Develop a Tailored Primary Care Quality Measure Set

ProMES is a comprehensive performance measure development approach firmly grounded in motivational theory and performance measurement.(2, 3) Through a facilitated focus-group based process, these measures are defined, weighted, and prioritized to create indicators of both overall effectiveness and specific aspects of daily work. This alignment helps individuals and teams to focus their effort more clearly on the most important aspects of their work (i.e., clinical performance) resulting in greater productivity, reduced stress, and less waste of effort.(2, 3) We utilize the ProMES definition of *productivity*, which is how effectively an organization uses its resources to achieve its goals.

Previous research in intensive care unit and mental health settings have shown successful use of ProMES to improve efficiency and quality of care. For example, in studies conducted with a German mental health hospital, the ProMES methodology was used across organizational levels: the top management team, samples of nurses, and three samples of technicians. As a whole, results indicated that overall productivity scores for participating units were, on average, .78 standard deviations higher after implementation.(3) Broader meta-analytic work examining the effectiveness of ProMES on improving productivity found a large overall effect size (δ = 1.44); with even higher effect sizes within particular subgroups (as high as δ = 2.21).

One unique feature of ProMES is the resulting measures include non-linear functions that explicitly tie performance levels on a given measure to its contribution to the organization’s values; in this way, the application of ProMES yields a more nuanced approach to prioritizing work than simple linear weights, while allowing direct comparison(s) between measure(s). In short, ProMES’ firm theoretical grounding, participatory development process, single index of productivity, and decades of research demonstrating its effectiveness differentiate it from other productivity enhancement and measurement programs and make it an ideal method for accomplishing the project’s objectives.

### Procedure

We adapted the ProMES steps involved in developing a performance measurement system to accomplish our first aim. Traditional ProMES methodology involves forming a design team of SMEs, developing objectives, generating performance indicators, and generating assessments of value for each indicator (a process called contingency development). Each step is described below. For this project, we adapted the indicator development process (Step 2) so that our design team selected from existing measures, rather than generating new ones. We identified potential indicators from an existing pool of primary care measures available on a national level through the VHA. Each potential indicator/existing measure was thoroughly vetted for quality against ProMES indicator quality criteria in order to mirror the methodologically rigorous characteristics of performance indicators *generated* through ProMES.

Step 1: Form Design Team

Our first step in adhering to the ProMES approach was the selection of our SME panel, known in ProMES parlance as a design team. We invited SMEs who met the following criteria: (1) filled gaps in the project team’s knowledge base, (2) possessed significant experience or a leadership role in primary care settings, and (3) were highly knowledgeable of and experienced with performance measures and/or electronic health record (EHR)-sourced data. To ensure representativeness in the design team, we additionally ensured the team consisted of experts from both inside and outside the VHA (our principal source of measure data) and represented multiple stakeholder groups including active primary care clinicians, administrators, performance measure data managers, and measurement experts. Our final team consisted of nine subject matter experts in addition to our project team.

Step 2: Identify Clinical Performance Objectives

A fundamental assumption of ProMES is that effective performance measurement cannot be accomplished without knowing what overarching performance objective is sought. Our design team was led by project team facilitators in a 1-day workshop to identify the clinical objectives for primary care (see Appendix A for the materials given to the design team). According to ProMES, objectives are defined as the essential things a unit (i.e., a primary care team) does to add value to the organization; in other words, objectives are the main result (and associated characteristics) of the primary care team’s work (e.g., amount, quality, timeliness). Examples of health care objectives include patient care performed according to quality standards, effective patient throughput, and personnel allocation matched to patient workload. The key question the design team was charged with answering was “what is the primary care team trying to accomplish when it delivers care?” The facilitators guided the design team to arrive at three to six objectives that met the following criteria: (1) was stated in clear terms, (2) designed so that if exactly that objective was accomplished, the facility (and thus the patient) would benefit; (3) the set of objectives cover all important aspects of clinical care; (4) was consistent with broader objectives (such as AHRQ conceptualizations of primary care); and (5) leadership was committed to each objective. Objectives were compared against these criteria to ensure their utility in facilitating the subsequent steps of the process (see Appendix A.1 for the Objectives Development Worksheet).

Step 3: Select Performance Indicators

*Selecting indicators.* Having identified primary care teams’ healthcare delivery objectives, the next step was to identify indicators for these objectives (see Appendix A.2 for the Indicator Selection Worksheet). In the classic ProMES process, these indicators would be generated *de novo*, rather than selected from a set. However, as such an abundance of measures already exist for primary care, the ProMES process was used slightly differently, yet innovatively – to cull, rather than generate, indicators of performance. For each objective, the design team answered the following question: “How would you show that the primary care team is meeting the stated objective?” To accomplish this, the design team received a document containing names, sources, and operational definitions of all outpatient clinical performance measures currently tracked by the VHA that were available at the team level. To ensure generalizability beyond the VHA health care system, any measures that captured VHA-specific processes or policies (e.g., homeless veteran measures; veteran patient portal usage) were excluded. Measures for the design team’s consideration were drawn from VHA’s principal outpatient clinical data sources and reports: VHA’s Corporate Data Warehouse, the principal repository of clinical and administrative data; the Strategic Analytics for Improvement and Learning (SAIL) system, VHA’s system for summarizing hospital performance, which includes the HEDIS measures used by most medical practices nationwide; and the Electronic Quality Measures (eQM), which provides, real-time, 100% sampling of patients for measure calculation.

The design team selected indicators via a two-step process. Design team members received documents describing the candidate measures two weeks in advance of the aforementioned day-long workshop so they would have adequate time for review. As part of the day-long workshop, the facilitators led the design team through a preliminary group voting process to identify the strongest candidate measures for each objective. In the weeks following the workshop, the team met virtually in a series of weekly 1 to 1½-hour facilitator-led meetings for up to a total of approximately 12 hours of meeting time. For each objective, the design team reviewed each candidate measure that survived the preliminary voting process. The team collectively narrowed down the list to a targeted set of 4-6 performance indicators per objective that captured the extent to which the primary care objectives were being achieved. Each indicator needed to meet multiple criteria regarding its validity, comprehensiveness, impact, feasibility, and usability (see Appendix A.2 for a complete list, compatible with National Quality Forum criteria).

Step 4: Prioritize Indicators by Developing Contingencies

*The value of contingencies.* Indicators provide information about what is valued in performance (e.g., the number of days between a fecal occult blood test (FOBT) order and a scheduled colonoscopy signals timeliness in care delivery); however, what level of performance is acceptable, or how much a given level of improvement is valued (Is an average of 7 days acceptable? How much worse is 8 days? 10? How much better is 5 days?) is provided by a ProMES tool called contingencies. One of the most novel aspects of ProMES is the use of contingencies. Contingency development generates a function for each indicator that shows how much the different amounts of the indicator (e.g., 5 vs. 7 days) contribute to overall effectiveness. By relating each indicator to overall effectiveness, they are put on the same measurement scale, which ranges from −100 to +100. Thus, the various indicators can be directly compared, prioritized, and combined into a single measure if needed. Most importantly, it reflects an explicit statement of what elements of primary care are valued, and what level of primary care-related performance is expected and valued by the team and the facility.

*Contingency curve examples.* Once the design team selected the indicators for each of the objectives, the classic ProMES approach was followed (see Pritchard et al., 2012 for details). In the classic ProMES approach, the design team determines three critical scores for each of the indicators: realistic minimum, realistic maximum, and the minimum level of acceptable performance. Figure 1 shows an example graph for an example indicator: percent of patients who missed appointments during the reporting period. In this example, the design team for the clinic would decide the realistic minimum (5% in this example) and the realistic maximum (25% in this example). Next, the minimum level of acceptable performance is determined. In this example, the minimum level of acceptable performance is when 15% of patients miss their appointments during the reporting period. The minimum level of acceptable performance corresponds to a score of zero on the effectiveness scale (see Figure 1).

**Figure 1.**
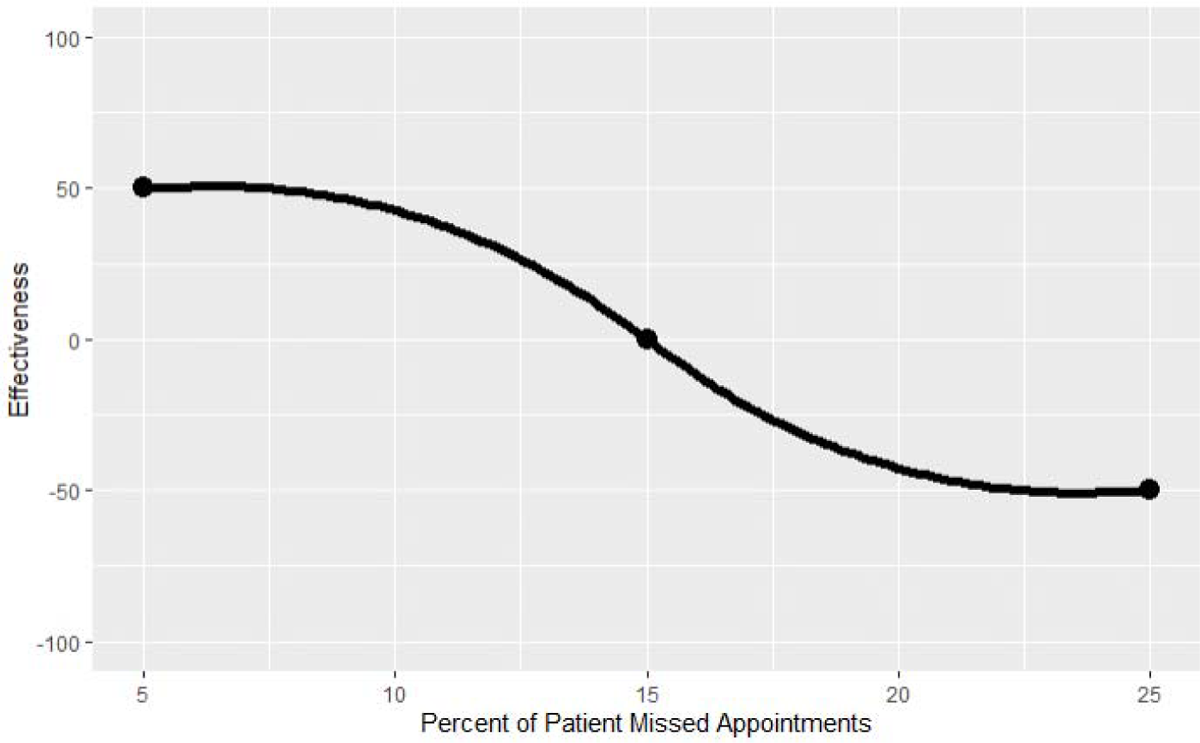
Example Contingency Curves

In the next step of the ProMES approach, the design team discusses and decides the effectiveness level that corresponds with the minimum and maximum indicator scores. The effectiveness score is the contribution the indicator makes to organizational goals. The fixed effectiveness scale range in ProMES is from −100 to +100. The most negative score, −100, means the indicator can have a large negative impact on the organization. On the other hand, the most positive impact an indicator can have on the organization is denoted by an effectiveness score of +100.(2, 16) As previously stated, an effectiveness score of zero is the minimum acceptable level of performance and does not harm or benefit the organization. Figure 1 shows that when the percent of patients decreases from 15% to 5%, the effectiveness score increases from 0 to +50. On the other hand, if the percent of patients increases from 15% to 25%, the effectiveness score decreases from 0 to −50. The larger an indicator’s effectiveness score range, the larger the impact it can have on the organization.(3, 16) In the example depicted in Figure 1, the indicator has an effectiveness score range from −50 to +50. Each indicator’s effectiveness range is compared to determine the relative importance of each indicator. Thus, this indicator’s range would then be compared with other indicators’ range to help prioritize which indicators impact the clinic the most.

It is also important to identify the appropriate curve shape, or contingency curve, for each indicator. As can be seen in Figures 2A and 2B, contingencies can be linear or a variant of a curve. A linear contingency means each change in the indicator level results in the same corresponding change in effectiveness,(3) which is not always appropriate; therefore, the design team identifies any points of nonlinearity. Figures 1, 2A, and 2B show a negative relationship between the indicator and effectiveness level, as the percent of patients increases, the effectiveness score decreases. However, the curve in Figure 1 shows a nonlinear relationship with an initial gradual decrease that has a steep decrease then continues to gradually decrease. In Figure 1, when the percent of patients goes from 10 to 5, there is an effectiveness gain of +12. However, if the percent of patient goes from 17 to 12, there is an effectiveness gain of about +60. In this example, the clinic would have more urgency in reducing the percent of patients from 17 to 12 than from 10 to 5. Figure 2A shows a linear relationship where each 5% change in the indicator results in a change of 25 in effectiveness. For example, if the percent of patients goes from 10 to 5, there is an effectiveness gain of +25. Figure 2B shows a curve with an initially steeper decrease then a gradual decrease followed by a steeper decrease. In Figure 2B, if the percent of patients goes from 10 to 5, there is an effectiveness gain of +38. However, if the percent of patients goes from 17 to 12, the effectiveness score increases by around 1. This suggests that for the example in Figure 2B, it is a higher priority to decrease percent of patients’ missed appointment from 10 to 5 percent. The design team determines the appropriate contingency curve for each indicator.

**Figure 2A.**
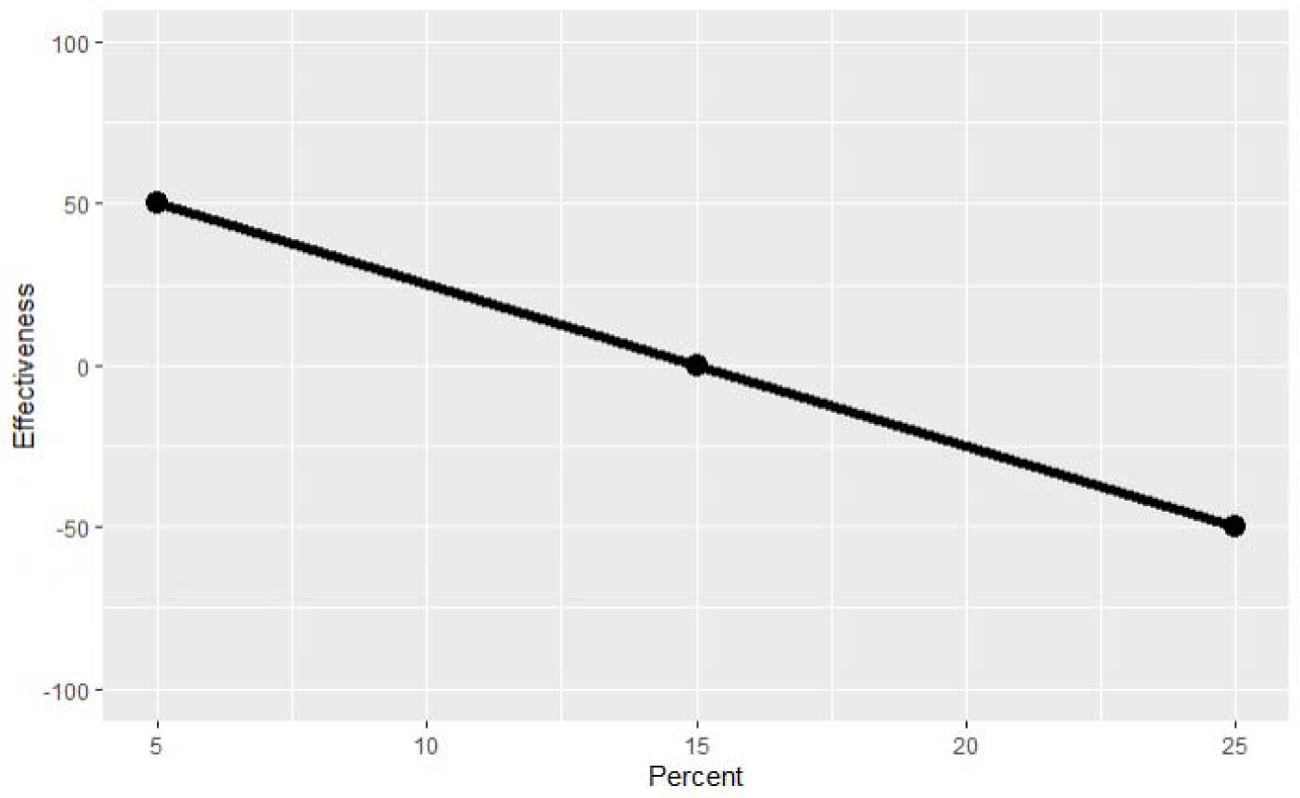
Example of a Linear Contingency

**Figure 2B.**
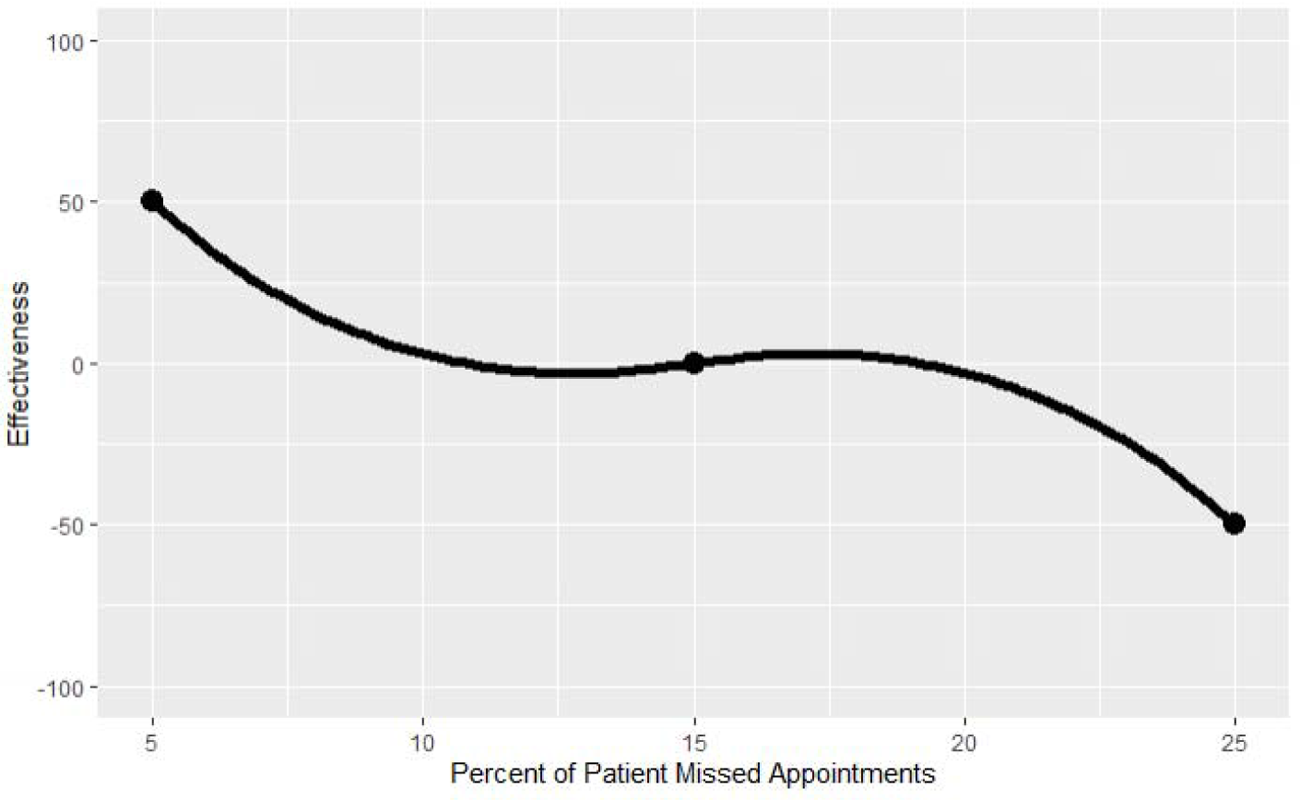
Example of a Possible Contingency Curve

*Creating contingencies.* The design team convened for a total of 6-8 hours (in multiple 1 to 1½-hour virtual meetings) to develop a contingency function for each indicator (see Appendix A.3 for the ProMES Contingency Development Worksheet). In doing so, members of the design team identified the maximum and minimum possible levels of performance, the minimum expected performance level, and scaled the various levels of performance to a common measure of effectiveness for each indicator (see Appendices B.4 and B.5 for the worksheets used, respectively). This was accomplished with the aid of the principal investigator serving as facilitator who, for each indicator, elicited discussion about each of the aforementioned data points, and guided each discussion to consensus through real-time, iterative polling of the design team. In sum, the creation of contingency curves serves to explicitly illustrate the importance and values of the organization’s policies.

## Results

### Objectives

The design team identified three fundamental objectives for delivery of high-quality primary care:

- *Objective 1*. Ensure patient has appropriate access to preventive, acute, or chronic health care services when needed.
- *Objective 2.* Build a trusting, effective, sustained partnership between the health-care team, the patient, and his/her caregiver(s) towards shared goals.
- *Objective 3.* Deliver safe and effective care that comprehensively addresses a given patient’s particular ecological, biological, and/or psychosocial needs.

A fourth objective, proactively monitor and engage with patient population(s) to optimize their treatment plans and health behaviors towards improved overall health, was originally identified. However, as the design team selected performance indicators for each of the objectives, it was evident that the fourth objective was subsumed under the other three objectives. Thus, the design team opted for a more succinct set of objectives and instead allowed for a slightly greater number of indicators in total, to capture this aspect of the primary care performance domain.

### Indicators and Contingency Curves

The design team selected sixteen performance indicators from the 44 pre-vetted measures that already exist in three different data sources for primary care, as outlined in Step 3. One indicator, Team 2 Day Post Discharge Contact Ratio, was selected as an indicator for both Objective 2 and 3. In addition, contingency curves were created for each of the indicators using the contingency functions developed by the design team as described in Step 4. Table 1 lists the primary care performance indicators selected by the design team, organized by objective. In Table 1, Team 2 Day Post Discharge Contact Ratio appears under both Objective 2 and 3. Please see Appendix B for detailed descriptions and contingency curves for each measure. Details of existing indicators by objective can be found in Appendix B.1.

**Table 1.**
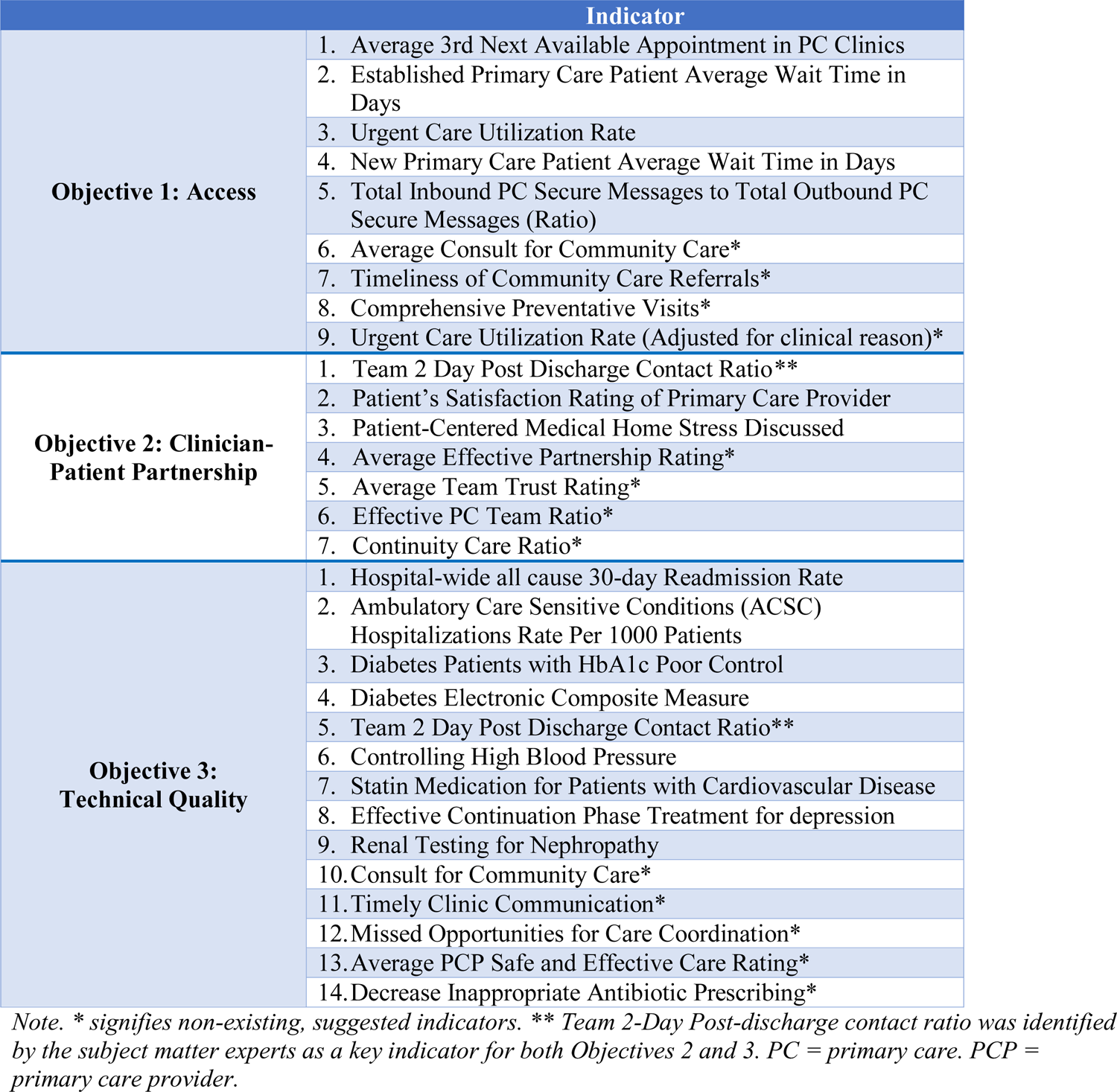
Consolidated, summary table of indicators including suggested, by objective

### Gaps in the Primary Care Performance Measurement Domain

The ProMES process is meant to guide measurement development or selection that yields a concise yet comprehensive list of indicators that contribute to an organization’s overall effectiveness.(3, 16) The design team identified areas of primary care performance that were not fully captured by the 16 existing indicators; therefore, the design team suggested additional indicators that are not currently measured but would bridge the criterion gaps. This gap was especially evident in Objective 2, where the design team was only able to select three relevant indicators from the existing measures. Table 1 lists these suggested indicators in brief; longer descriptions of suggested indicators can be found in Appendix B.2.

## Discussion

This white paper details the application of an evidence-based approach to produce a curated set of primary care quality indicators, prioritized according to their overall contribution and value to primary care. Our design team of nationally recognized SMEs joined together with diverse stakeholder groups in a national panel to collectively identify three primary care delivery objectives, 16 currently existing indicators of primary care quality, and 13 indicators requiring further development, to address current gaps in the primary care performance measurement domain. The selected indicators were selected independent of clinic size or configuration, so that clinics of many configurations (e.g., public vs. private, large vs. small, rural vs urban, team-based vs. traditional) could benefit from their use.

Results speak to the type of indicators readily available for each stated objective of care. Our first care objective, centered around access to care, highlights the overall importance of primary care facilities to maintain sufficient capacity. In robustly assessing this care objective, indicator(s) for this objective also capture(s) insufficient or ineffective access to primary care by capturing patients’ visit(s) to the emergency department instead of their primary care clinic for their care. Interestingly, our second care objective, covering the importance of maintaining a trusting partnership with the patient, contained relatively few existing indicators, primarily due to the challenges inherent in self-report data and lack of available objective measures for processes of care delivery face to face (e.g., building rapport). Finally, our third objective of achieving technical quality contains an abundance of measures, reflecting the condition-specific origins of the performance measurement movement in health care. Measures for these objectives revolve around evidence-based processes for the treatment of highly prevalent chronic physical and mental conditions most commonly managed in the primary care setting; similar to the access objective, an additional facet captures ineffective technical quality through hospitalizations resulting from ambulatory care sensitive conditions and hospital admissions. In turn, our study revealed the existing discrepancies between ideal and current measurement of care quality.

Specifically, although assessing the quality of less tangible but essential activities such as disease education and shared decision-making in the context of relationship building with patients would ideally form a significant proportion of quality care measurement, a disproportionate number of existing measures focused on the more readily measured technical competencies in clinical practice (Objective 3). A balanced number of indicators across all three objectives would be a better comprehensive set of care quality measures.

### Implications

Performance measures selected as part of our modified ProMES process assist in the implementation of targeted care quality measures prioritized in accordance to their value in primary care. Although many efforts exist to assess technical proficiency and patient-related outcomes, lack of guidance in their implementation, interpretation, and use as proxies for care quality may not directly assess the effectiveness of primary care teams or practices at meeting their objectives. By deriving high-value metrics, organized by objective with quantifiable prioritization, we anticipate the results of this paper will apply to a diverse set of stakeholders, including but not limited to policy-makers, primary care clinicians, and administrators in healthcare organizations. As more primary care settings transition toward a team-based delivery approach, we believe our curated set of indicators provide more meaningful, appropriate, and high-quality assessments of primary care team effectiveness, enabling low-resource settings (such as Federally Qualified Health Clinics [FQHCs]) to improve the care they deliver.

Our findings bear implications to the scientific and primary care communities. Practically, our proposed measure set establishes precedent for communicating policy explicitly within care quality measurements. Namely, in regard to care quality among primary care clinics, our targeted manner of measurement explicitly prioritizes the value of each care measure in their overall contribution and value to primary care. In practice, results of using these measures could translate to greater actionability for quality improvement initiatives in the primary care setting. Further, the proposed measures could be adopted to evaluate clinics who wish to pursue PCMH certification through the Joint Commission. Based on currently available data collection techniques, this set of measures offer a less obtrusive, and thus less resource-intensive manner by which team-based primary care quality can be ascertained in that it relies on existing sources of quality data and methods of data capture. Theoretically, our measures and their respective objectives provide clear domains for measurement of primary care quality, which could lead to quality improvement interventions that are better aligned with the goals and the promise of primary care.

### Limitations

Our resulting set of measures is necessarily limited by what was available in existing VHA data sources; indeed, such limits were observed in the need to propose new measures that did not exist in the current data source in order to comprehensively cover the primary care performance domain. Thus, in the current measure set outcome measures are overrepresented relative to process measures. This is also evident in the reverse pattern presenting in the proposed measures relative to the existing ones.

Although leveraging available measures avoids several unintended consequences involved in care quality measure creation, existing data sources have several documented problems. First, use of electronic health record data for research purposes is often questioned due to completeness and quality of data entered. Second, sampling strategies used to collect more subjective measures of care quality (e.g., patient satisfaction) collect data from a sub-set of a clinical teams’ patient panel and may not fully represent the team’s provision of care.

While no data source is perfect, we hope to target efforts on improving the quality of health systems by building upon existing techniques rather than inventing new methods of data capture.

Furthermore, not all practices or systems have the infrastructure needed to measure, capture, and utilize the data to measure care quality. Some practices or systems may not have the resources or have competing resource demands, such as needing to invest resources in equipment or buildings rather than performance measurement. Funding initiatives for practices that want to adopt a PCMH model should provide sufficient incentives to significantly reform infrastructure including a measurement system to measure care quality.

A core assumption of this paper is that the primary care performance objectives result in patients’ improved health and that they align with patients’ goals. Moreover, we are mindful of patient centered issues and needs that are not captured by the objectives, such as the healing comfort a healthcare professional can provide. Future efforts should more explicitly capture the needs of the patient when identifying clinical performance objectives and indicators.

Finally, psychometric procedures have not been conducted on the selected measures as a set to confirm statistically that the selected indicators indeed load onto their respective objectives. Obviously, this check cannot be performed on the proposed measures that do not, as of yet, exist.

### Conclusions and Future Directions

We conclude that through the use of an evidence-based approach to performance measure development, a targeted set of performance measures can be curated to track progress toward specific objectives in delivering high-quality primary care. Our measure set provides an actionable catalogue of measures that can serve as a first step toward interoperability of electronic health record systems. Future work toward this goal should address both logistical considerations (e.g., data capture, common data/programming language) and lingering measurement challenges, such as the best way to operationalize these measures for teams working in complex and shifting situations (e.g., rotating team members).

## Data Availability

Data for this paper will be available upon written request to the corresponding author.

## Appendix A: Performance Indicator Quality Criteria

The materials on the following pages were used to guide the expert panel convened for the process of selecting the primary care quality measures.

## Appendix A.1: Objectives Development Worksheet

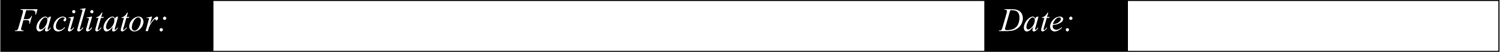

***Facilitator Instructions:*** *Follow the instructions in Pritchard et al. 2011 for conducting the focus groups. Pay special attention to ensure the criteria for quality objectives are met (see criteria at the end of this worksheet) as you work with the expert panel. Use this worksheet to capture the final objectives identified by the expert panel, as well as their rationale for why they are relevant to primary care, including detail about how each one meets the criteria. Add more boxes as necessary to capture objectives; however, the number of objectives should be manageable, normally 3 to 8*.

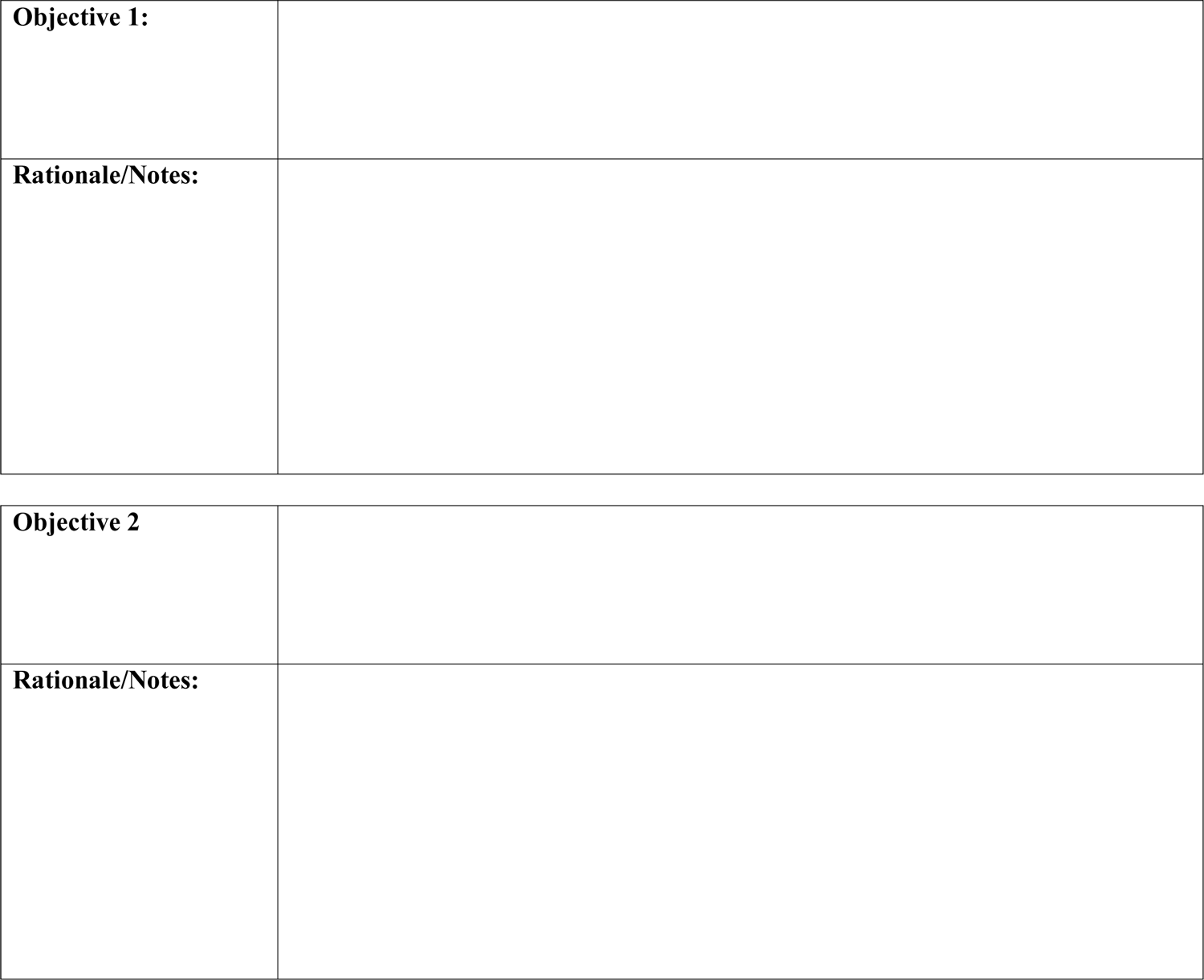

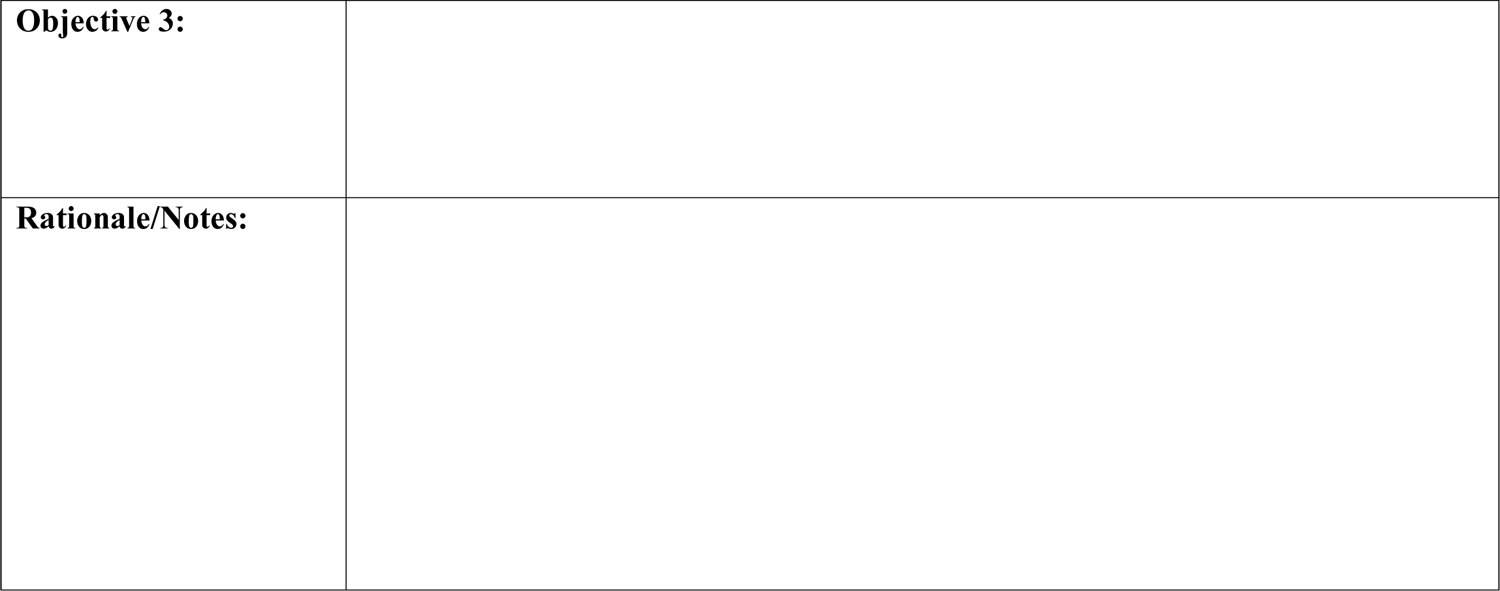

**Rationale/Notes:**
|  |
| --- |
| **Objective 3:** |
| **Rationale/Notes:** |

**Criteria for ProMES Objectives**

Below are the criteria used to evaluate the quality of performance objectives according to the ProMES model. Objectives that meet these criteria are important prerequisites for identifying and developing appropriate performance measures.

- Objectives should be stated in clear terms
- Objectives should be designed so that if exactly that objective was accomplished, the organization would benefit
- The *set* of objectives must cover all important aspects of the work (in our case, all important aspects of primary care)
- Objectives must be consistent with the objectives of the broader organization
- Leadership must be committed to *each* objective

## Appendix A.2: Indicator Selection Worksheet

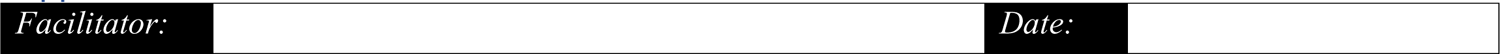

***Facilitator Instructions:*** *Follow the instructions in Pritchard et al. 2011 for conducting the focus groups. Note that this step is different from classic ProMES in that the expert panel will be selecting from existing measures (sample measures are provided in Table 1 for your reference) rather than developing measures* de novo*. Pay special attention to ensure the criteria for quality indicators are met (see criteria at the end of this worksheet) as you work with the expert panel. Use this worksheet to capture the final indicators identified by the expert panel, as well as their rationale for why they are relevant to primary care, including detail about how each one meets the criteria. Add more boxes as necessary to capture indicators; however, the number of objectives should be manageable, normally 5-15 total, with at least one but no more than 5 or 6 per objective*.

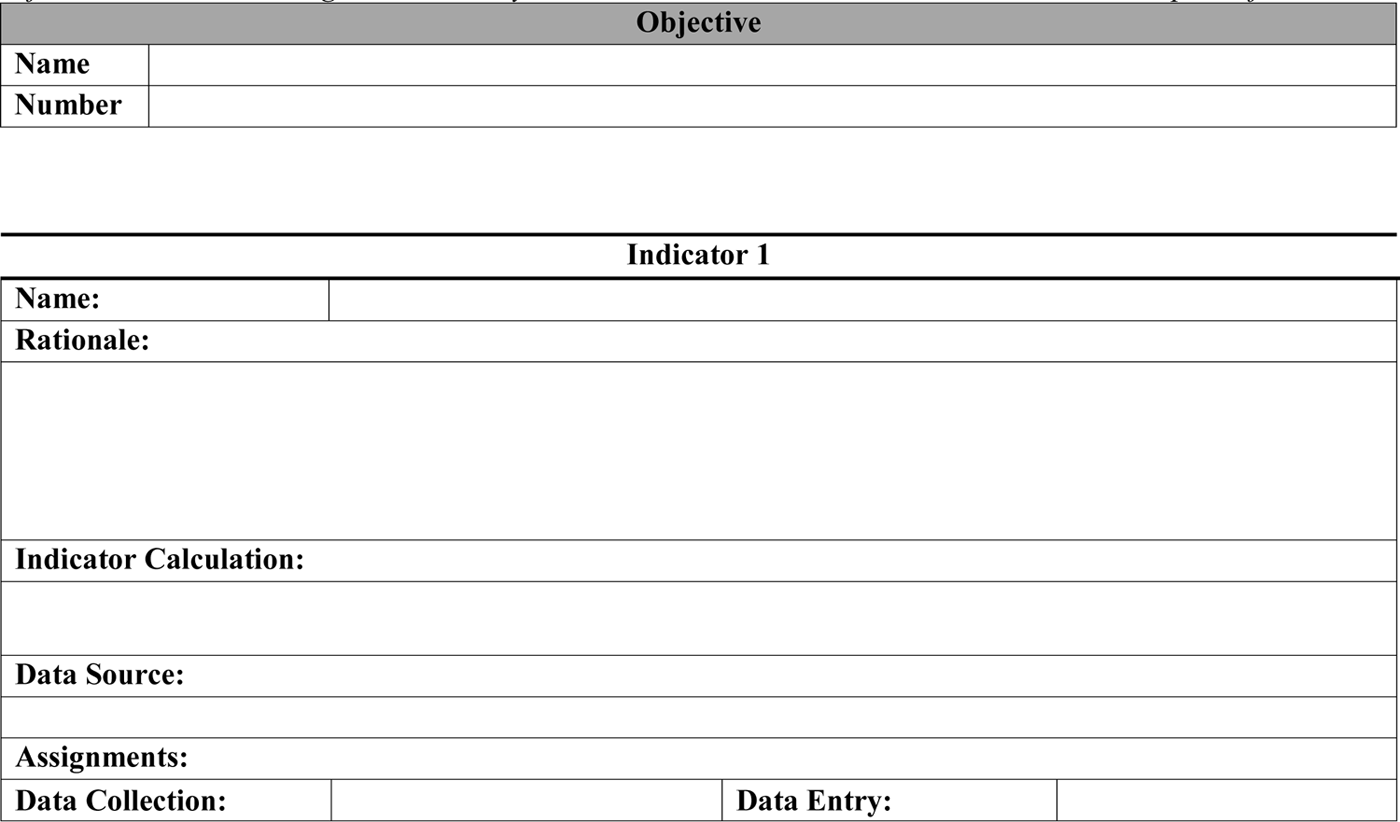

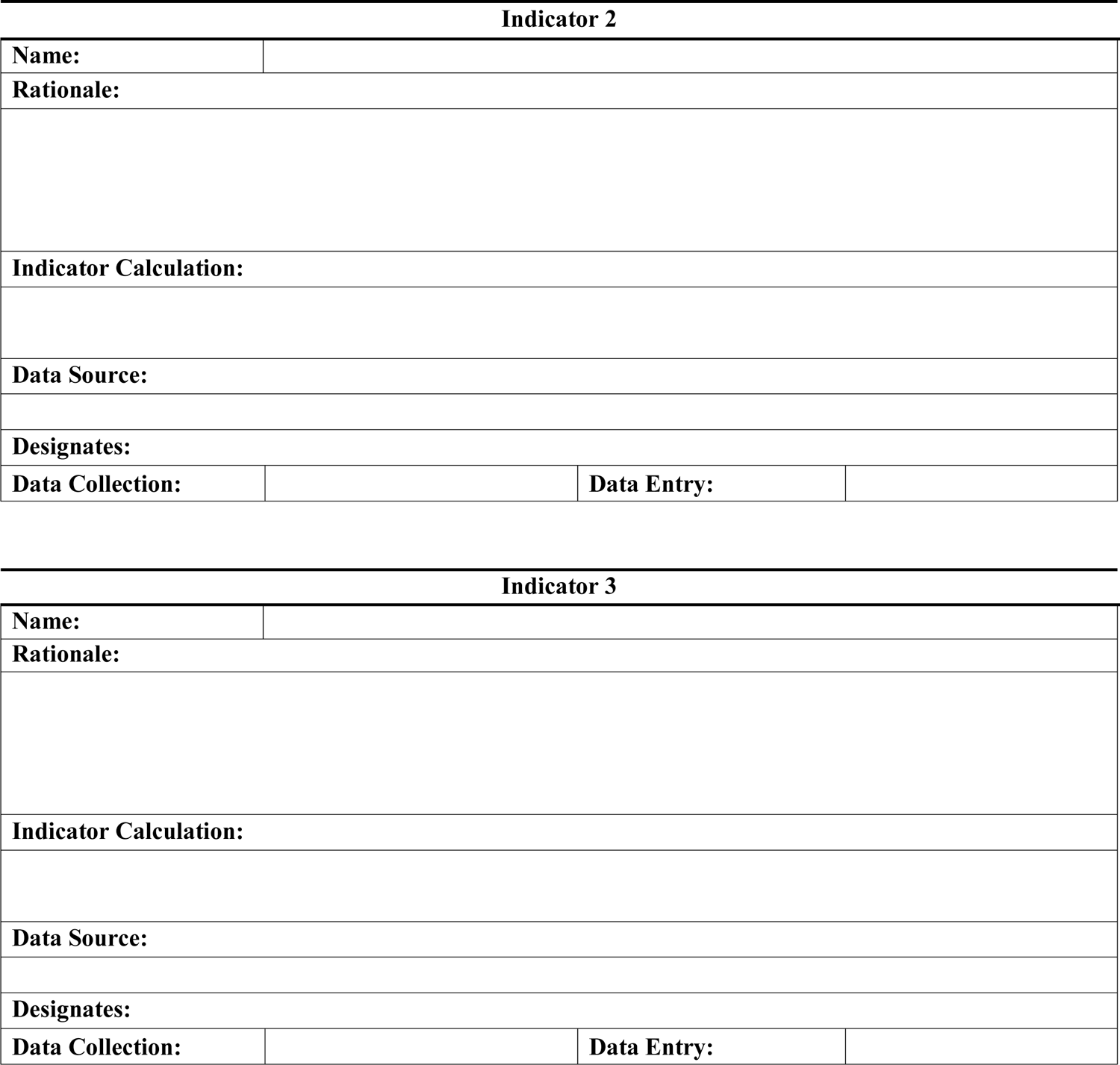

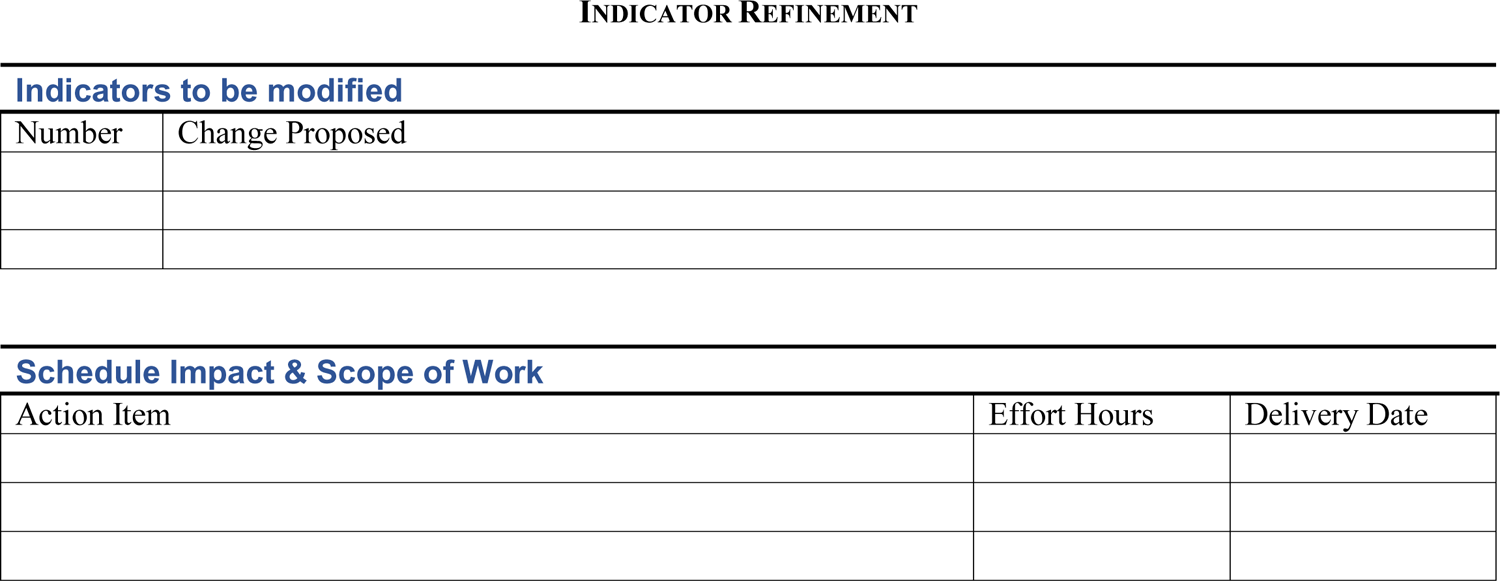

**Criteria for ProMES Indicators**

Below are the criteria used to evaluate the quality of a performance measure according to the ProMES model. For consistency with more health-care specific model, they are organized according to the criteria used by the National Quality Forum^46^ to evaluate clinical measures.

Validity/Reliability

- Indicators must validly measure the objective
- Indicators must be largely under the control of unit personnel
- The information provided by the indicator must be neither too general nor too specific.

Comprehensiveness (not part of NQF criteria)

- All important aspects of each objective must be covered by the *set* of indicators

Impact (Value)

- Indicators must be consistent with the objectives of the broader organization
- Indicators should be designed so that if the indicator was maximized (i.e., perfect score), the organization would benefit (value – similar to NQF’s Impact)

Feasibility

- Leadership must be committed to *each* indicator
- Accurate indicator data must be cost effective to collect

Usability

- Indicators must be understandable and meaningful to unit personnel
- It must be possible to provide information on the indicator in a timely manner

## Appendix A.3: ProMES Contingency Development Worksheet

***Facilitator Instructions:*** *Follow the instructions in Pritchard et al. 2011 for conducting the focus groups. For each indicator, the design team must accomplish the following:*

1. *<u>Identify the maximum and minimum possible values for each indicator.</u>* What is the absolute minimum possible turnaround time to schedule an appointment once the ordering provider places a referral request? Zero days? 1 Day? What is the maximum possible turnaround time? If the answer is an infinite value, (e.g., the referral appointment never gets placed at all) an unacceptably large number can be used (e.g., 120 days).
2. *<u>Identify the minimum expected performance level</u>.* What is the minimum expected turnaround time for scheduling an appointment (i.e., in this case, the greatest number of days it can take and still be considered acceptable)? This should be the expected standard, that is, a level of performance that gains you neither reprimand nor praise.
3. *Rank each indicator in value to the facility if each indicator was at its maximum, and assign effectiveness levels.* The design team will rank the indicator maximums in terms of overall importance to the team’s objectives. Whichever indicator is believed to make the greatest contribution is given the rank of 1, and its maximum value is given an effectiveness value of +100. All other indicators’ maximum values for a given objective, which have been ranked in order of contribution, are then given values on the effectiveness scale relative to the indicator with the rank of one. For example, if the maximum of a given indicator was only half as important to the coordination effectiveness of the team as the most important maximum, they would give it a value of +50.
4. *Rank each indicator in value to the facility if each indicator was at its minimum, and assign effectiveness levels.* A similar process as that described in step 3 above is conducted with the minimum values for each indicator. The design team will rank the indicator minimums in terms of overall importance to the team’s coordination work. Each minimum is first ranked as to which would be the worst for the team if all indicators were at the zero points and one was at its minimum. The worst minimum is then compared to the best maximum in order to decide its effectiveness value; the idea is to compare how bad the worst negative is to how good the best positive is, as it is not safe to assume that the worst possible performance is equally bad as the best possible performance is good. For example, the most negative minimum might be seen as only 80% as bad as the best positive indicator value is good. All other indicators’ minimum values for a given objective, which have been ranked in order of contribution, are then given values on the effectiveness scale relative to the indicator with the rank of one. For example, if the minimum of a given indicator was only half as important to the coordination effectiveness of the PACT as the most detrimental minimum, and the most detrimental minimum had a value of −50, they would give it a value of −25.
5. *<u>Plot basic contingency function and fill in remaining points in the plot.</u>* The design team will then, for each indicator, decide the effectiveness of each value in the function. For example, if the minimum expected turnaround time for scheduling referral appointments is seven days (effectiveness of 0), and the best possible turnaround time is 24 hours, how much value to coordination does a turnaround time of 3 days garner, relative to 24 hours? Half as much? 80% as much (i.e., getting the turnaround time to down to 3 days is almost as good as 24 hours)? If the latter, then a turnaround of 3 days would receive an effectiveness score of +80. This process is repeated with all major values on the scale in order to more accurately define the shape of the contingency curve for each indicator. The shape of the plot does not have to be (and in fact is often unlikely to be) linear. A sample contingency function is provided for your reference in Figure 1.

**Figure 1.**
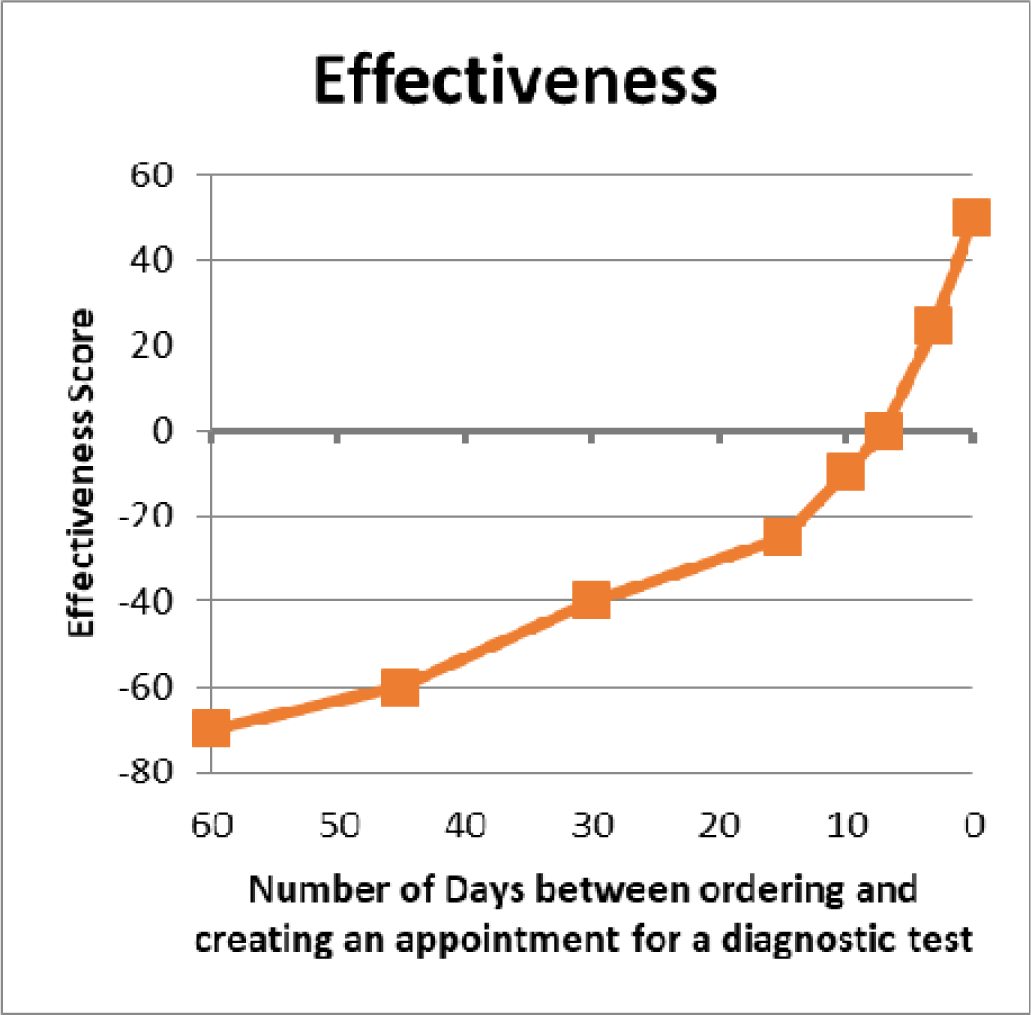
Sample Contingency Function.

**Table 1.**
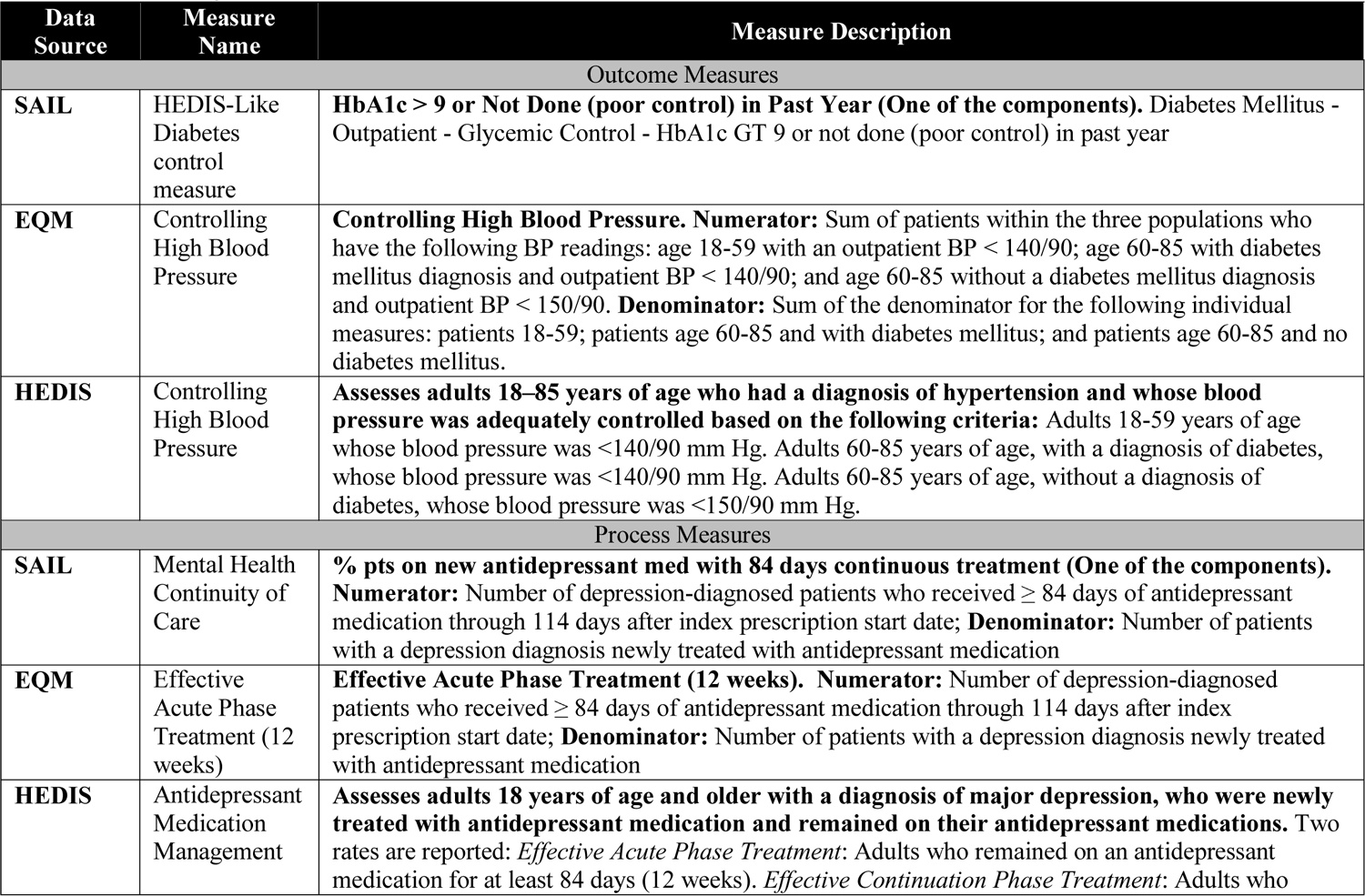
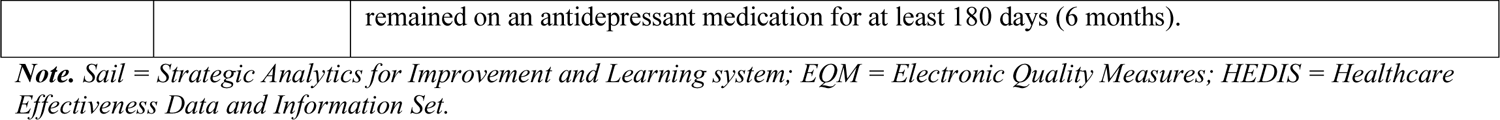
Sample Indicators

## Appendix A.4: Contingency Function Values Worksheet

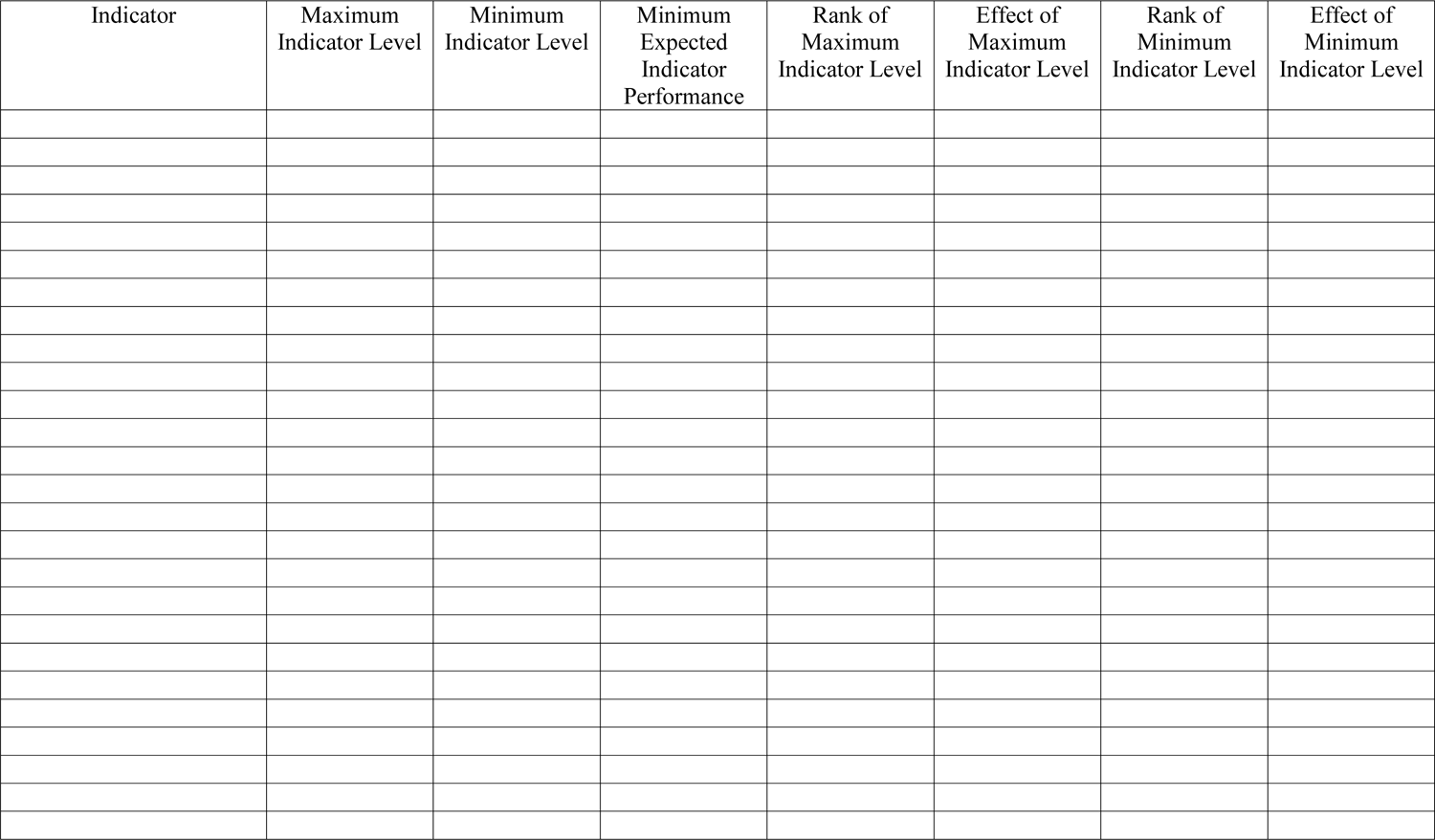

## Appendix A.5: Contingency Plot Worksheet

***Facilitator Instructions****: Use one chart per indicator to plot the contingency function*.

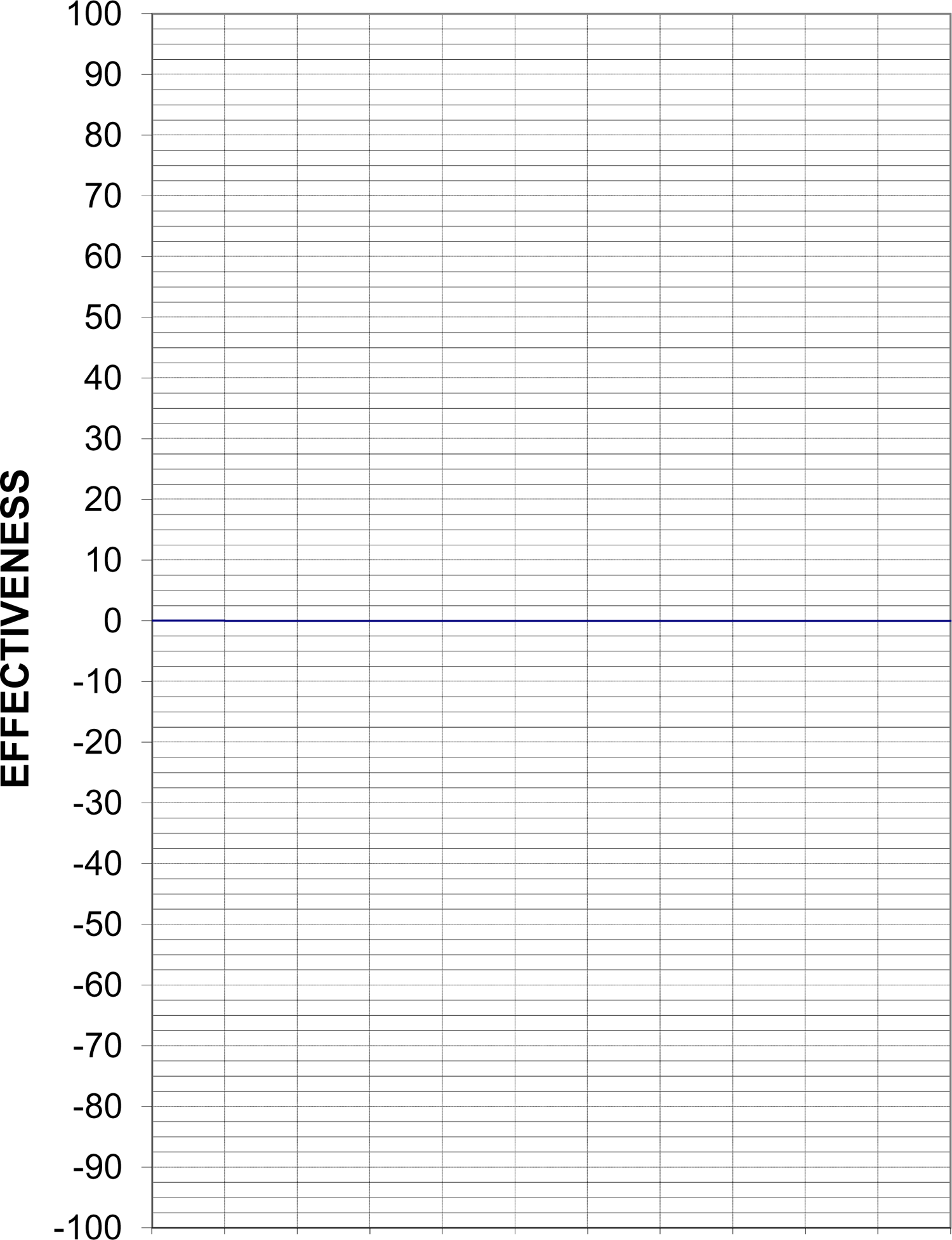

## Appendix B: Catalog of Primary Care Quality Measures

## Appendix B.1: Detail of Existing Indicators by Objective

*Indicator 1. Average 3^rd^ Next Available Appointment in PC Clinics*

Description. The average waiting time in days between a completed appointment and the 3^rd^ Next Available Appointment slot for each primary care clinic. A snapshot is taken on the first day of each month for the prior month’s activity. The wait times in days until the 3^rd^ Next Available Appointment is averaged for completed appointments.

Contingency curve.

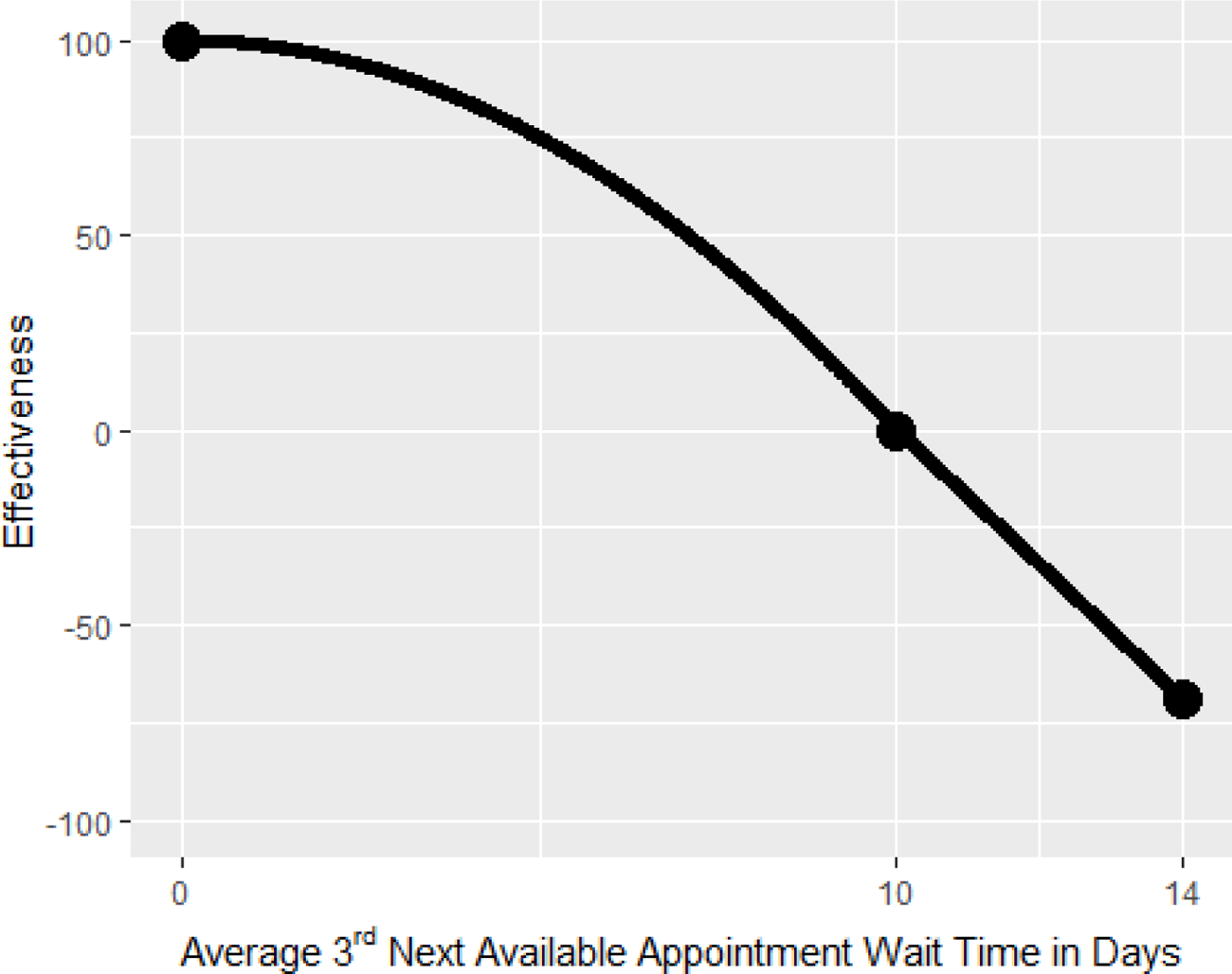

Interpretation. The graph shows that when patients in PC clinics wait on average 10 days for the 3^rd^ Next Available Appointment, the effectiveness level is 0; this means the minimum performance is met. If the wait time decreases to 0 days, then the effectiveness score increases to 100. On the other hand, if the wait time increases to 14 days, the effectiveness score decreases to about −70.

Data source. PACT Compass

Mnemonic. Average 3^rd^ Next Available in PC Clinics (322, 323, 350)

*Indicator 2. Established Primary Care Patient Average Wait Time in Days (Excluding Compensation & Pension appointments)*

Description. The average number of calendar days between an established patient’s PC completed appointment and earliest of three possible preferred (desired) dates from the completed appointment date.

Contingency curve.

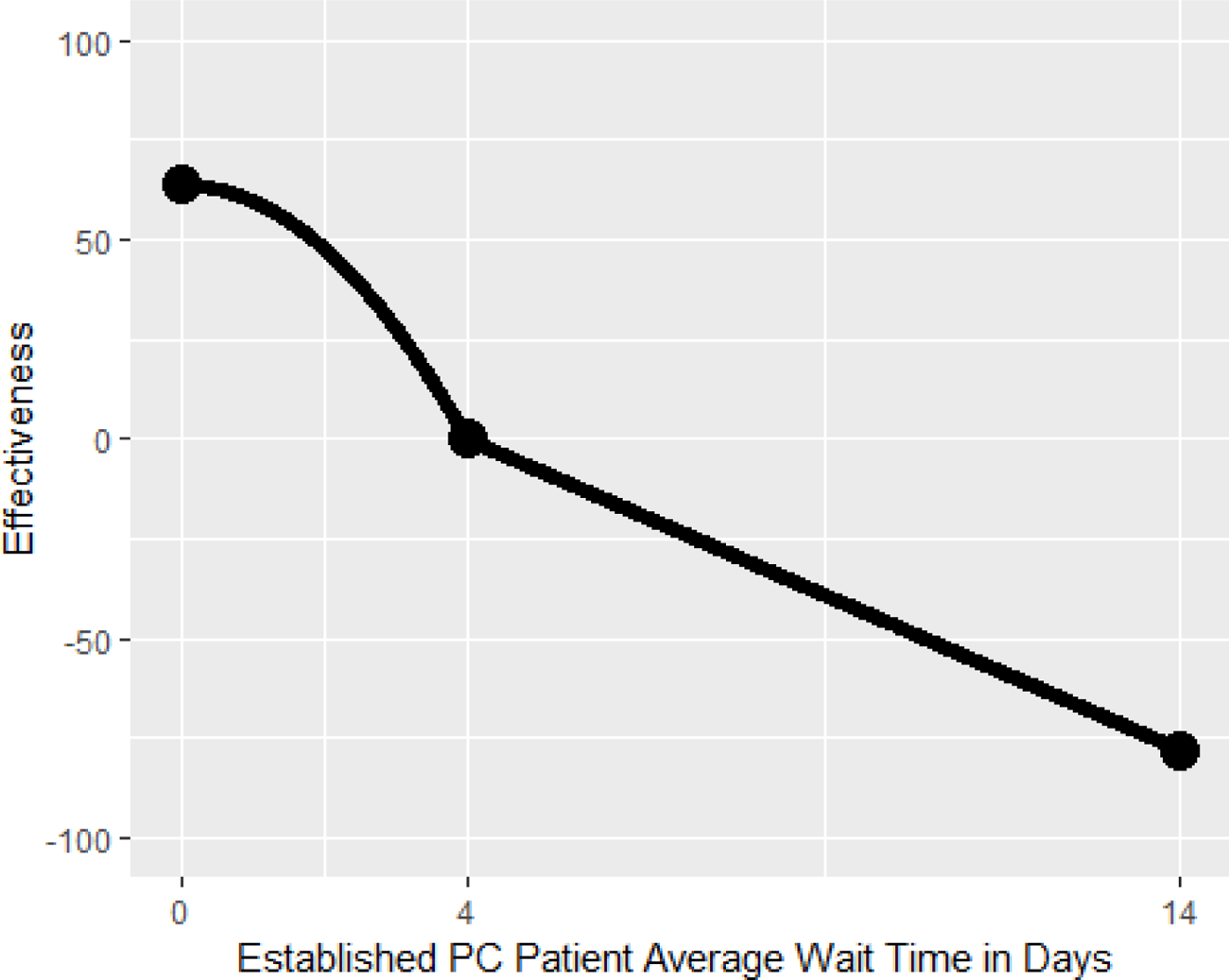

Interpretation. The horizontal axis shows the average wait time in days between an established patient’s PC appointment and the earliest of three possible dates (electronic wait list, cancelled by clinic appointment, and completed appointment). The graph shows that when the average wait time for an established PC patient is 4 days, the effectiveness level is 0; this means the minimum performance is met. If the average wait time decreases to 0 days, then the effectiveness score increases to about 70. On the other hand, if the average wait time increases to 14 days, the effectiveness score decreases to about −75.

Data source. PACT Compass

Mnemonic. Established PC Patient Average Wait Time in Days (Excluding Compensation & Pension)

*Indicator 3. New Primary Care Patient Average Wait Time in Days (Excluding Compensation & Pension appointments)*

Description. The average number of calendar days between a new patient’s PC completed appointment and the earliest of three possible preferred (desired) dates from the completed appointment date.

Contingency curve.

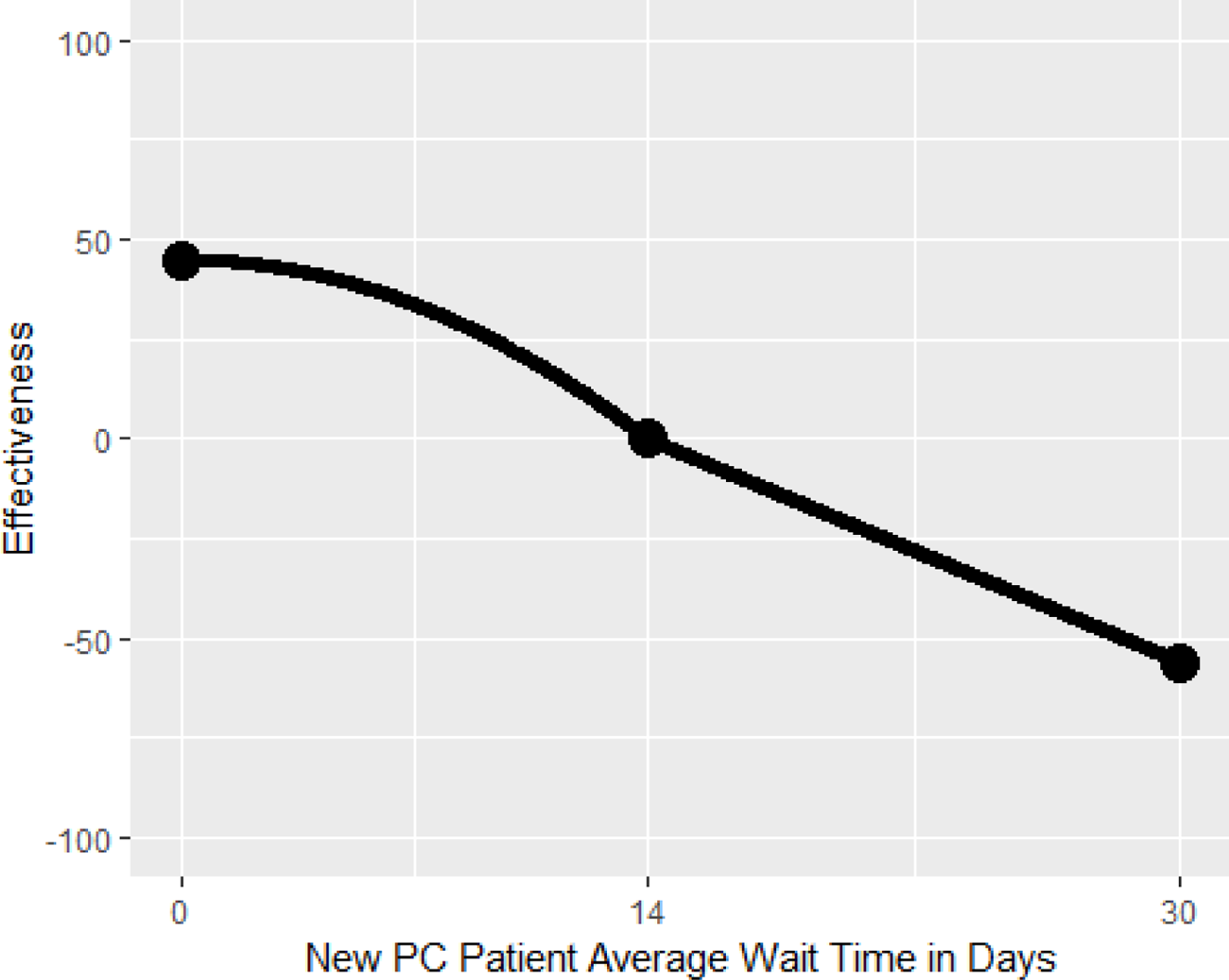

Interpretation. The horizontal axis shows the average wait time in days between a new PC patient’s completed appointment and the earliest of three possible preferred dates (electronic wait list, cancelled by clinic appointment, and completed appointment). When the average wait time for a new PC patient is 14 days, the effectiveness level is 0; this means the minimum performance is met. If the average wait time decreases to 0 days, then the effectiveness score increases to almost 50. On the other hand, if the average new PC patient wait time increases to 30 days, the effectiveness score decreases to about −55.

Data source. PACT Compass

Mnemonic. New PC Patient Wait Time in Days (Excluding Compensation & Pension)

*Indicator 4. Total Inbound PC Secure Messages to Total Outbound PC Secure Messages (Ratio)* Description. This measure is a ratio representing the total number of secure messages sent by a patient assigned to a given primary care team divided by the total number of secure messages sent from a primary care team member to a patient assigned to that primary care team during the reporting period.

Contingency curve.

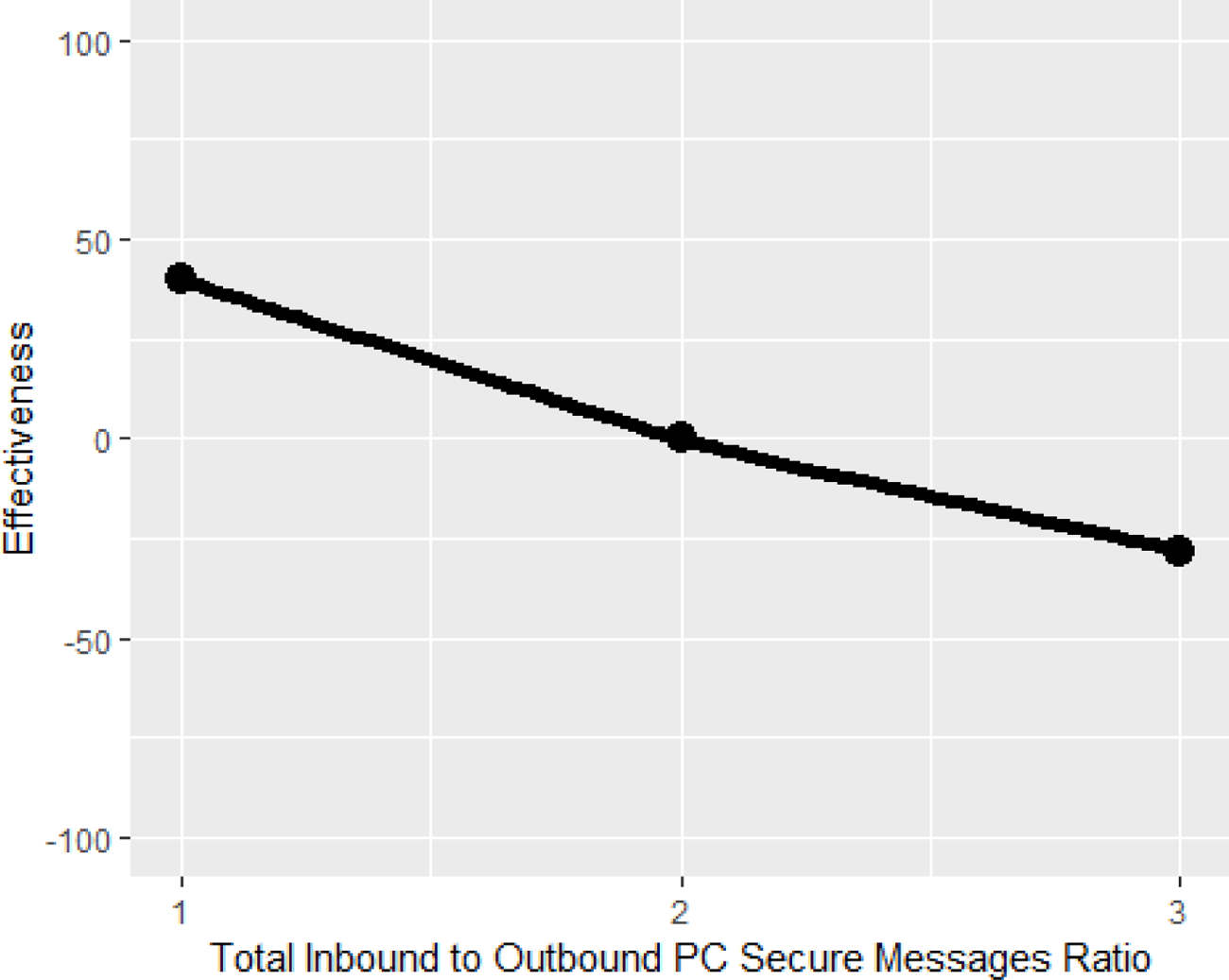

Interpretation. The graph shows that when the total inbound PC secure messages to total outbound PC secure messages ratio is 2, the effectiveness level is 0; this means the minimum performance is met. If the ratio decreases to 1, then the effectiveness score increases to about 45. On the other hand, if the total inbound PC secure messages to total outbound PC secure messages ratio increases to 3, the effectiveness score decreases to about −25.

Data source. PACT Compass

Mnemonic. Inbound PC Secure Messages/ Outbound PC Secure Messages (Ratio)

*Indicator 5. Urgent Care Utilization Rate*

Description. The total number of ER/Urgent Care encounters for assigned primary care patients in the last 12 months divided by the team assignments.

Contingency curve.

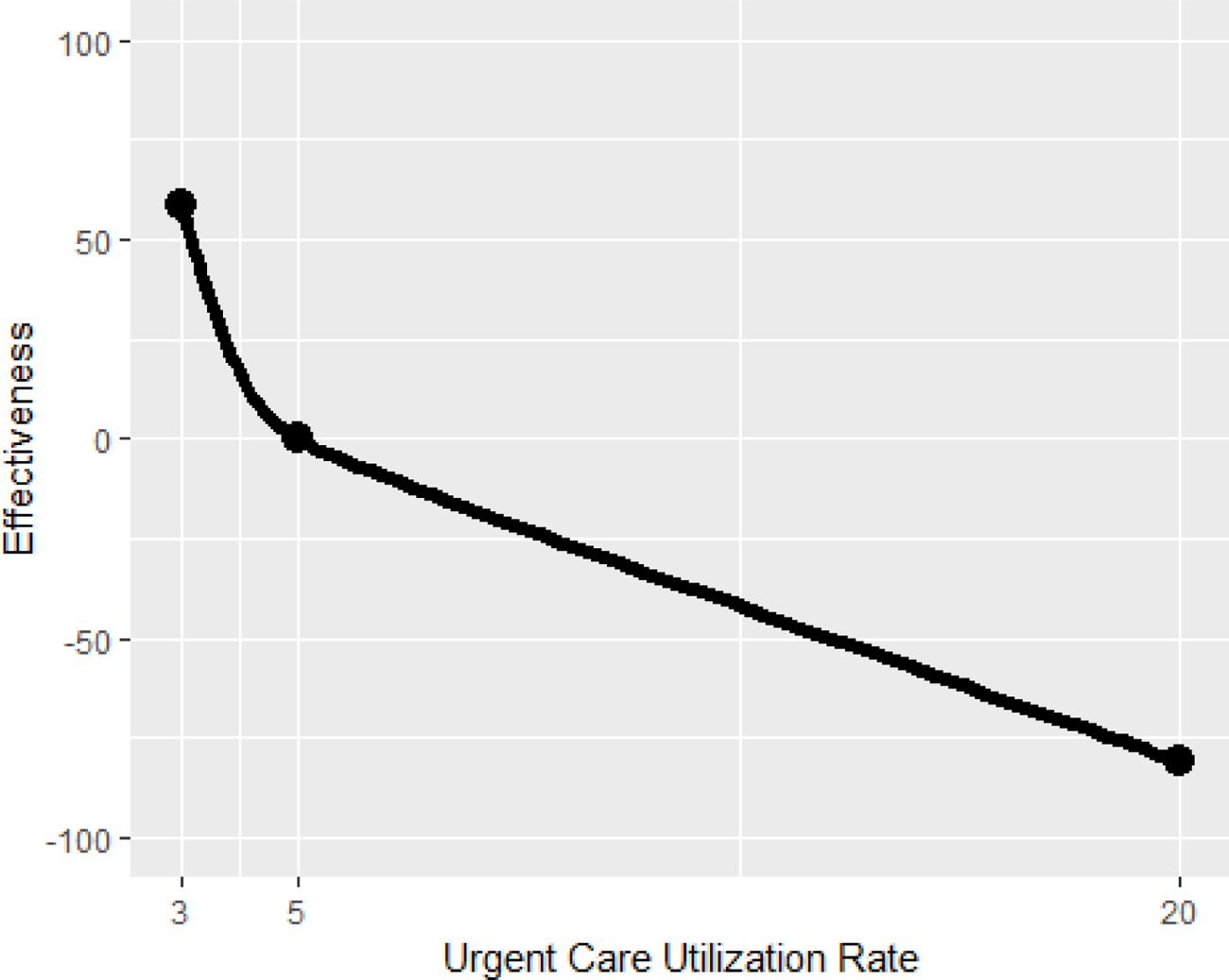

Interpretation. The graph shows that when patients’ urgent care utilization rate per team is 5, the effectiveness level is 0; this means the minimum performance is met. If the urgent care utilization rate decreases to 3, then the effectiveness score increases to about 60. On the other hand, if the urgent care utilization rate increases to 20, the effectiveness score decreases to about −80.

Data source. PACT Compass

Mnemonic. Urgent Care Utilization Rate

Objective 2. Build a trusting, effective, sustained partnership between the health-care team, the patient, and his/her caregiver(s) towards shared goals.

*Indicator 1. Team 2 Day Post Discharge Contact Ratio*

Description. This measure represents the percentage of patients assigned to a given primary care team who were contacted within two days of being discharged (DC) from inpatient care. The post discharge contact is only counted if the individual contacting the patient has a team role of administrative associate, care manager, clinical associate, designated women’s health primary care provider, clinical pharmacist, physician-attending, or primary care provider. Patients are excluded from this measure if they are discharged from an observation specialty and/or are readmitted within two business days to any healthcare facility.

*Note. This measure is also an indicator of safe and effective care delivery and also appears under Objective 3*.

Contingency curve.

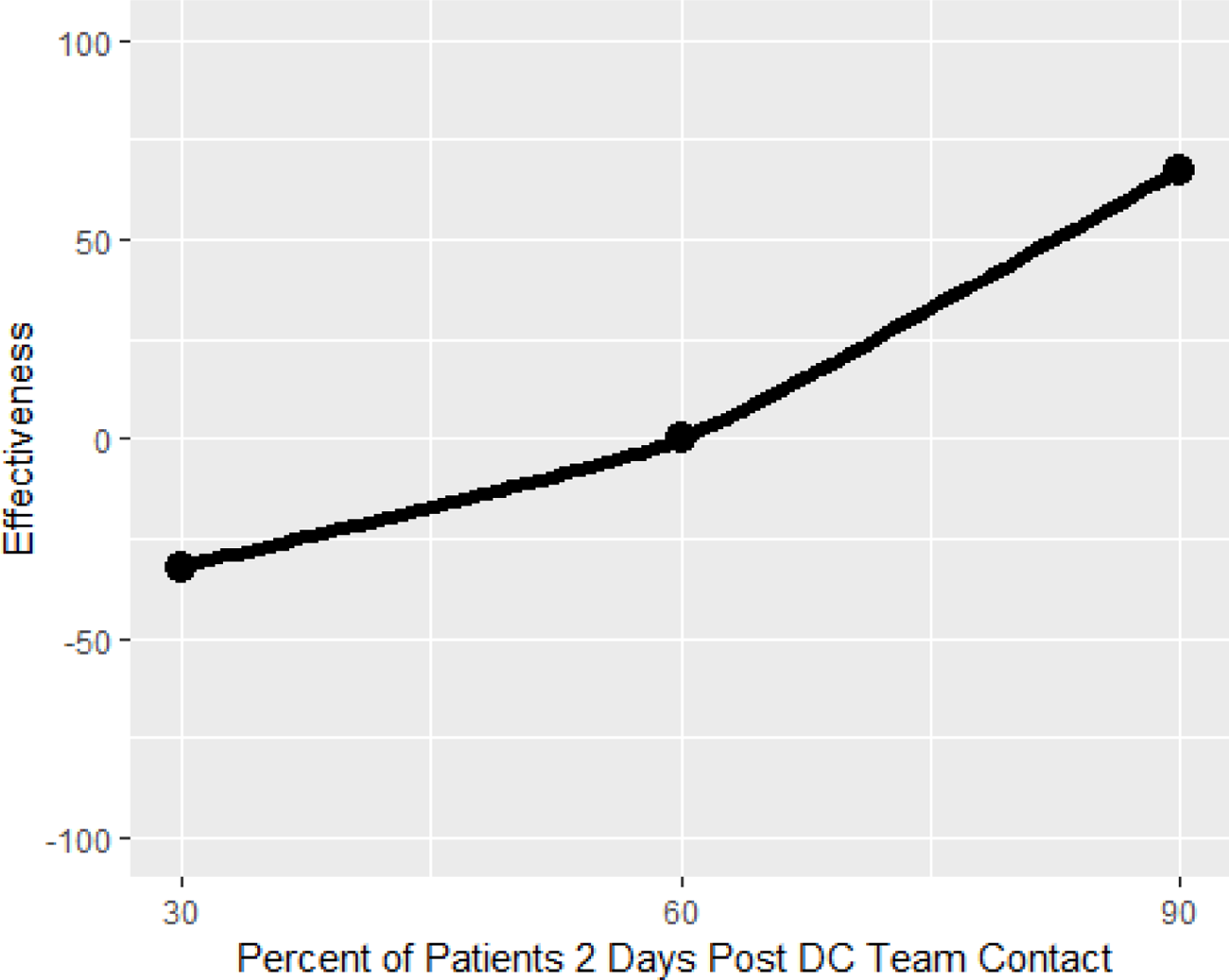

Interpretation. The graph shows that when the percent of patients contacted within two days of being discharged from inpatient care is 60, the effectiveness level is 0; this means the minimum performance is met. If the contact ratio decreases to 30, then the effectiveness score decreases to about −30. On the other hand, if the contact ratio increases to 90, the effectiveness score increases to about 70.

Data source. PACT Compass

Mnemonic. Team 2Day Post DC Contact Ratio – Core Teamlet Only

*Indicator 2. Patient’s Satisfaction Rating of Primary Care Provider*

Description. This measure is based on a question which asks patients to rate their primary care provider, after a qualifying visit on a 0 to 10 scale with 0 being the worst possible and 10 being the best. This measure is the percentage of patients’ responses of a 9 or 10 (the top two categories).

Contingency curve.

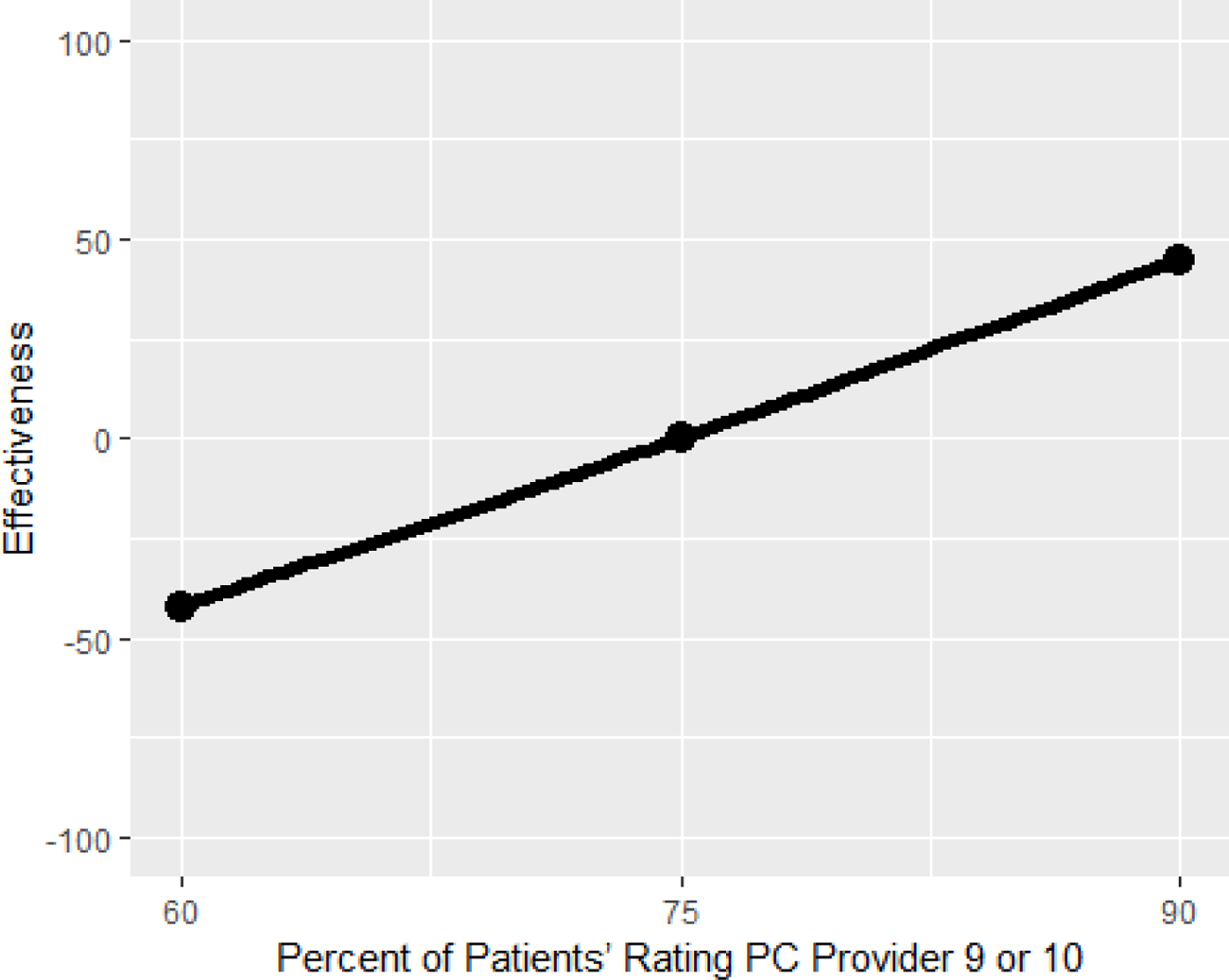

Interpretation. The graph shows that when the percentage of patients rating their provider 9 or 10 is 75, the effectiveness level is 0; this means the minimum performance is met. If the percentage of patients rating their provider 9 or 10 decreases to 60, then the effectiveness score decreases to about −45. On the other hand, if the percentage increases to 90, the effectiveness score increases to about 45.

Data source. SHEP

Mnemonic. Rating of Primary Care Provider (Q32) PCMH Survey

*Indicator 3. Patient-Centered Medical Home Stress Discussed*

Description. This measure comes from a question which asks the patient if, “in the last 6 months, did anyone in this provider’s office talk to you about things in your life that worry you or cause you stress”? The measure reflects the percentage of patients who responded “yes” to the question.

Contingency curve.

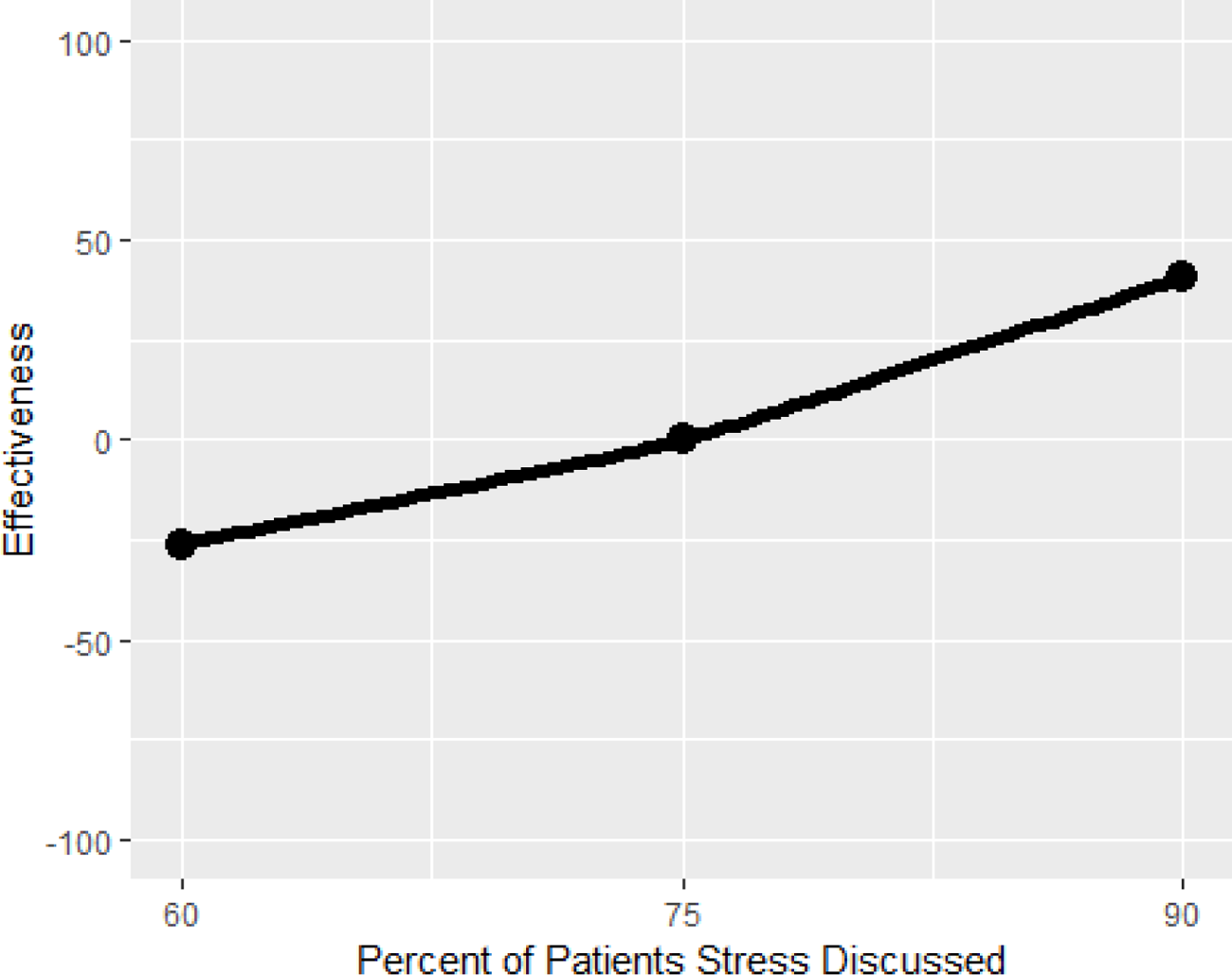

Interpretation. The graph shows that when the percentage of patients that discussed their stress with someone in the provider’s office is 75, the effectiveness level is 0; this means the minimum performance is met. If the percentage of patients that discussed their stress decreases to 60, then the effectiveness score decreases to −25. On the other hand, if the percentage of patients that discussed their stress increases to 90, the effectiveness score increases to about 45.

Data Source. SHEP

Mnemonic. PCMH Stress Discussed (Q40)

Objective 3: Deliver safe and effective care that comprehensively addresses a given patient’s particular ecological, biological, and/or psychosocial needs.

*Indicator 1. Hospital-wide all cause 30-day Readmission Rate*

Description. Rate of unplanned readmissions in the 30 days after discharge from a hospitalization. Rate is derived from a composite of five statistical models, built from groups of hospitalizations that are clinically related: Cardiorespiratory, Cardiovascular, Medicine, Neurology, and Surgery/Gyn. The measure does not count planned readmission. This measure is designed to provide aggregate and detailed views of the data to assist managers and clinicians in identifying potential gaps when transitioning patients through different stages of the recovery processes.

Contingency curve.

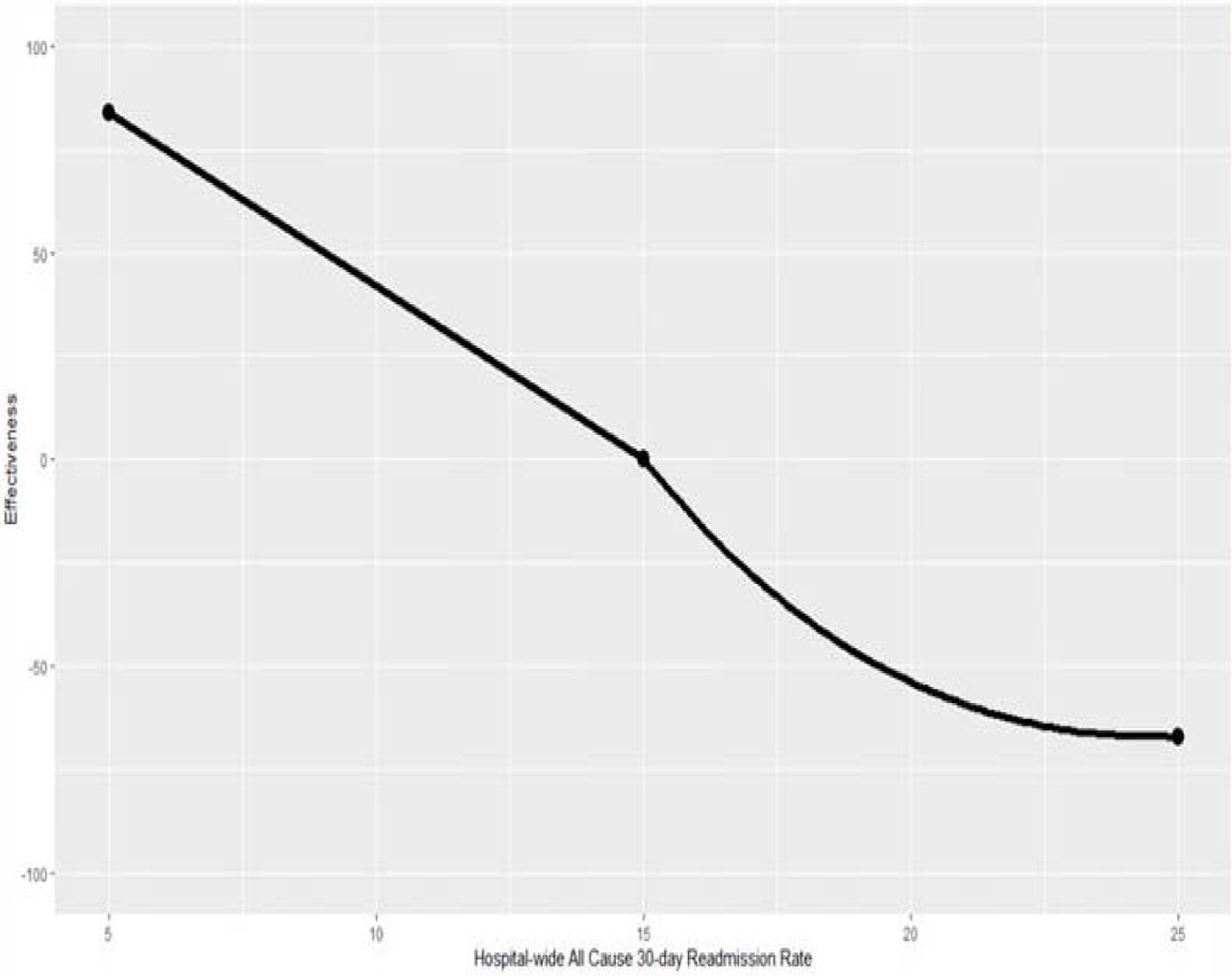

Interpretation. The graph shows that when the hospital-wide all cause 30-day readmission rate is 15, the effectiveness level is 0; this means the minimum performance is met. If the hospital-wide all cause 30-d y readmission rate decreases to 5, then the effectiveness score increases to about 85. On the other hand, if the rate increases to 25, the effectiveness score decreases to about −65.

Data Source. SAIL

Mnemonic. Hospital-wide All Cause 30-day Readmission Rate

*Indicator 2. Ambulatory Care Sensitive Conditions (ACSC) Hospitalizations Rate Per 1000 Patients* Description. Hospitalizations due to ACSCs such as hypertension, congestive heart failure, and pneumonia can typically be avoidable or preventable if ambulatory care is provided in a timely and effective manner. It has been well established that effective primary care is associated with lower hospitalization due to ACSCs. This rate is calculated by AHRQ using state population and the equation is ACSC hospitalizations divided by ACSC population. A similar option to calculate the ACSC hospitalization rate per 1,000 patients is by calculating the number of inpatients with a principal diagnosis of ACSC divided by the number of total patients with any diagnosis of ACSC. See Appendix C for an example of the data used to calculate ACSC hospitalization rate per 1,000 patients. This equation can be adapted to the team level by using patient panel size instead of state population.

Contingency curve.

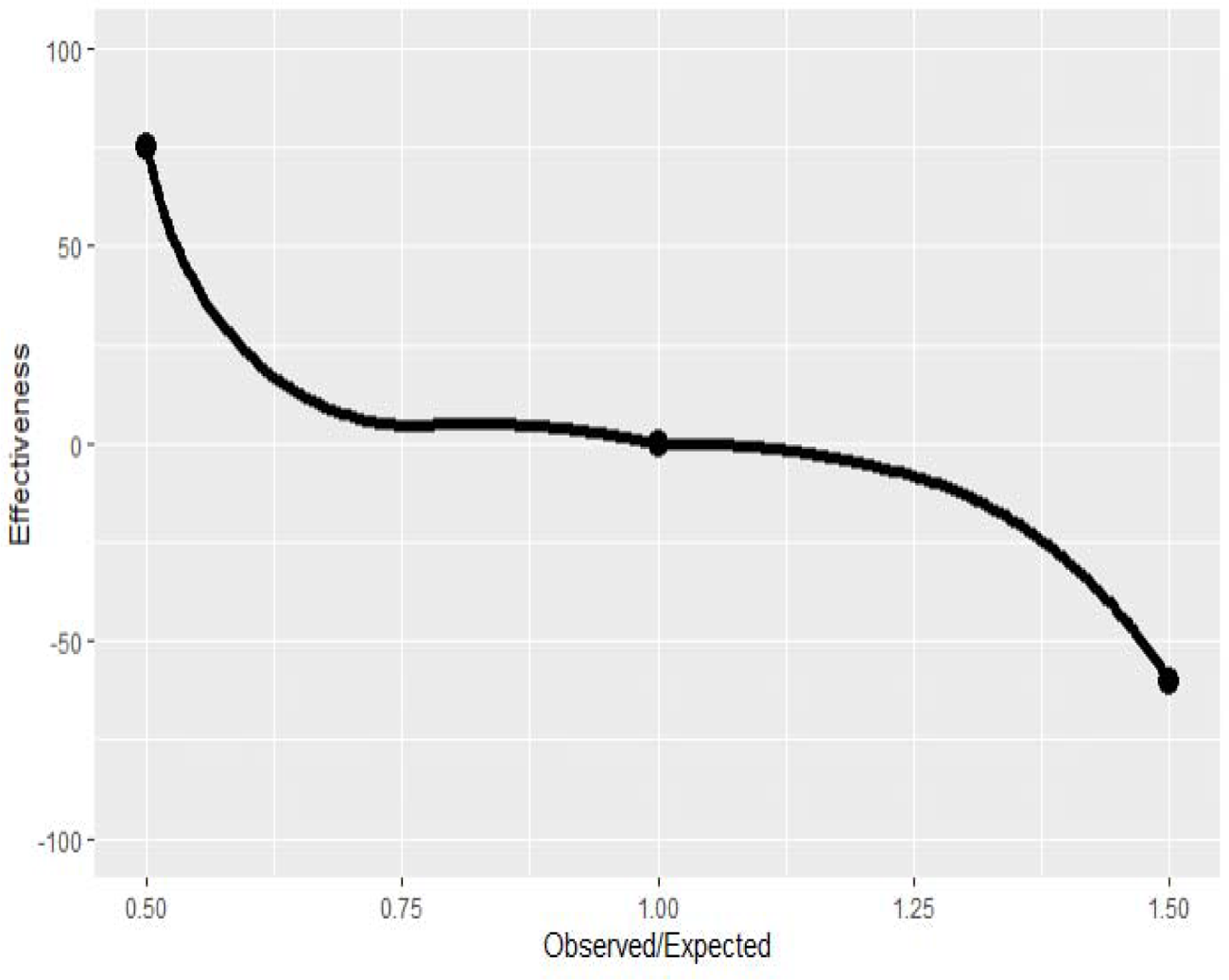

Interpretation. The graph above shows that when the rate of observed over expected ACSC hospitalizations per 1000 is 1, the effectiveness level is 0; this means the minimum performance is met. If the rate of observed over expected ACSC hospitalizations per 1000 decreases to .5, then the effectiveness score increases to 75. On the other hand, if the rate of observed over expected ACSC hospitalizations per 1000 increases to 1.5, the effectiveness score decreases to about −60.

Data source. SAIL

Mnemonic. ACSC (Ambulatory Care Sensitive Conditions) Hospitalizations

*Indicator 3. Diabetes Patients with HbA1c Poor Control*

Description. This measure represents the number of patients diagnosed with diabetes mellitus between the ages of 18 and 75 whose HbA1c score is greater than 9 or who show no evidence of having their HbA1c tested within the last year.

Contingency curve.

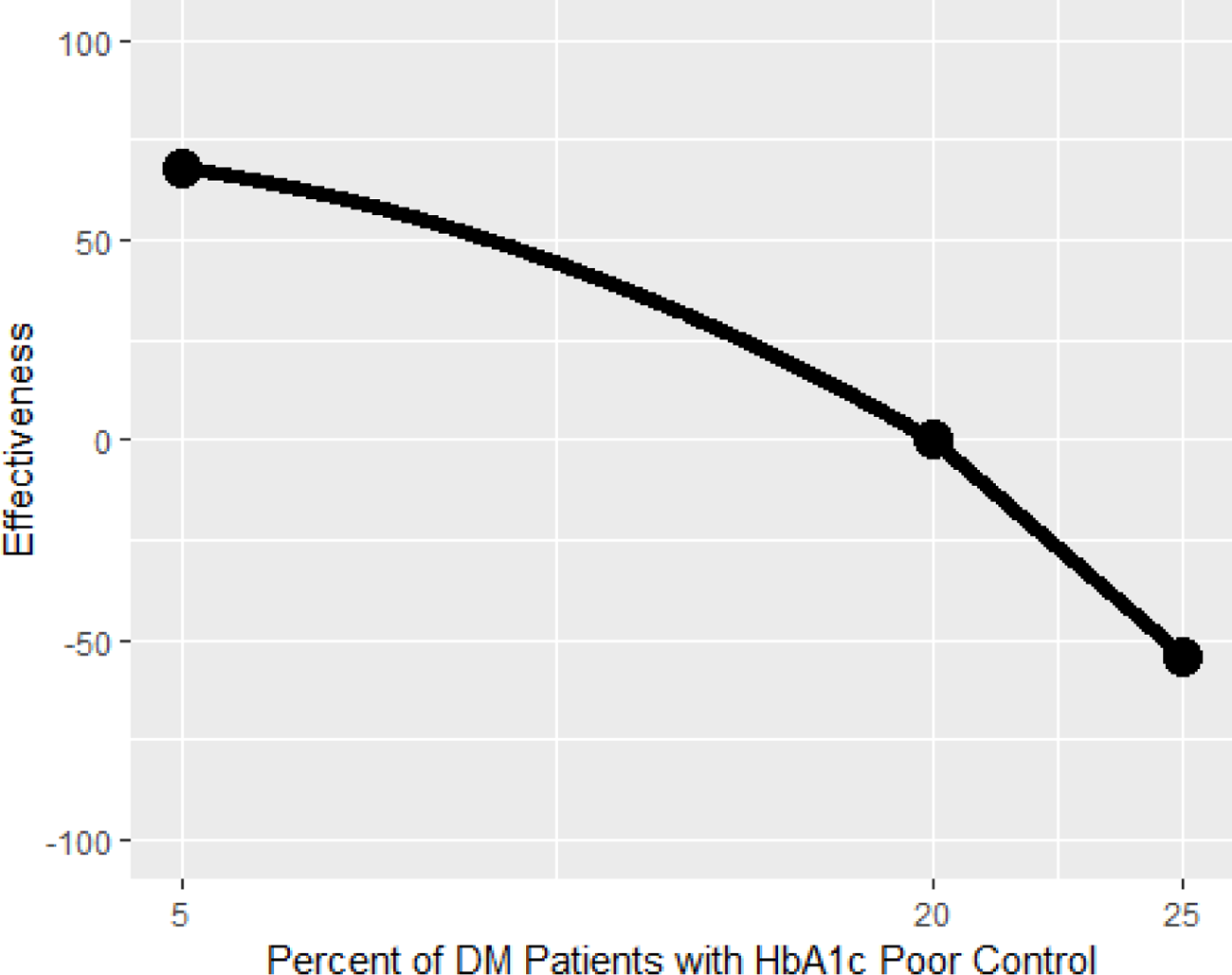

Interpretation. The graph shows that when the percentage of patients diagnosed with diabetes mellitus with HbA1c poor control is 20, the effectiveness level is 0; this means the minimum performance is met. If the percentage of diabetes patients with HbA1c poor control decreases to 5, then the effectiveness score increases to about 70. On the other hand, if the percentage of diabetes patients with HbA1c poor control increases to 25, the effectiveness score decreases to about −55.

Data source. eQM

Mnemonic. dmg23h_ec

*Indicator 4. Diabetes Electronic Composite Measure*

Description. This measure is a composite of the “Diabetes Patients with HbA1c Poor Control” measure and the HEDIS measure “Diabetes Mellitus—Outpatient: HbA1c Annual Testing” which is the number of patients between 18 and 75 years of age who have had HbA1c testing within the measurement year.

Contingency curve.

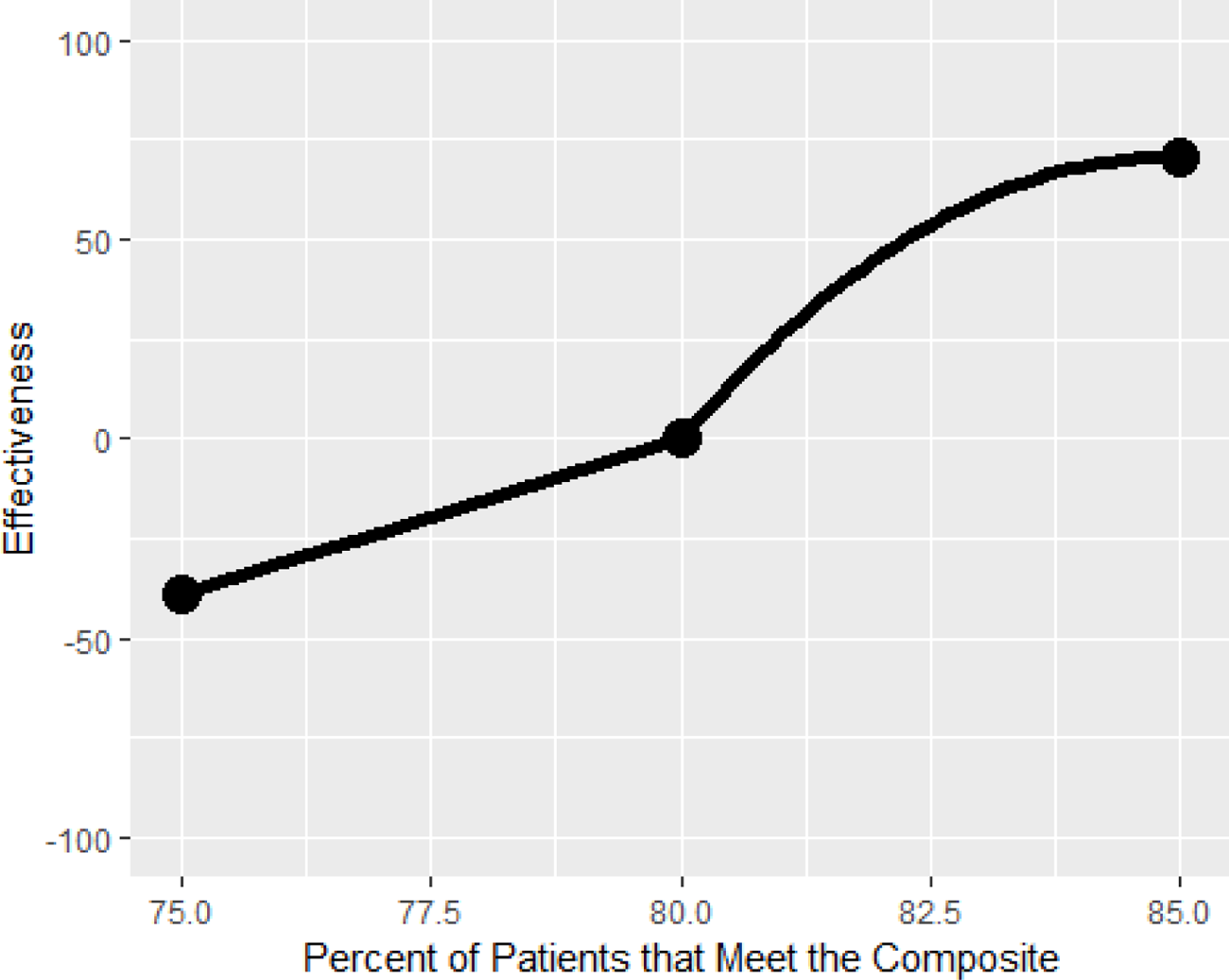

Interpretation. The graph shows that when the percent of patients that meet the composite is 80, the effectiveness level is 0; this means the minimum performance is met. If the percent of patients that meet the electronic composite decreases to 75, then the effectiveness score decreases to about −40. On the other hand, if the percent of patients that meet the composite increases to 85, the effectiveness score increases to about 70.

Data source. eQM

Mnemonic. dmg90_ec

*Indicator 5. Controlling High Blood Pressure*

Description. This is the number of patients between the ages of 18 and 85 with a diagnosis of hypertension within the first six months of the measurement year who are later found to have: + A blood pressure of less than 140/90 for outpatient patients aged 18-59; + A blood pressure of less than 140/90 for outpatients aged 60-85 with a diagnosis of diabetes mellitus (DM); Or +A blood pressure of less than 150/90 for outpatient patients aged 60-85 without a DM diagnosis.

Contingency curve.

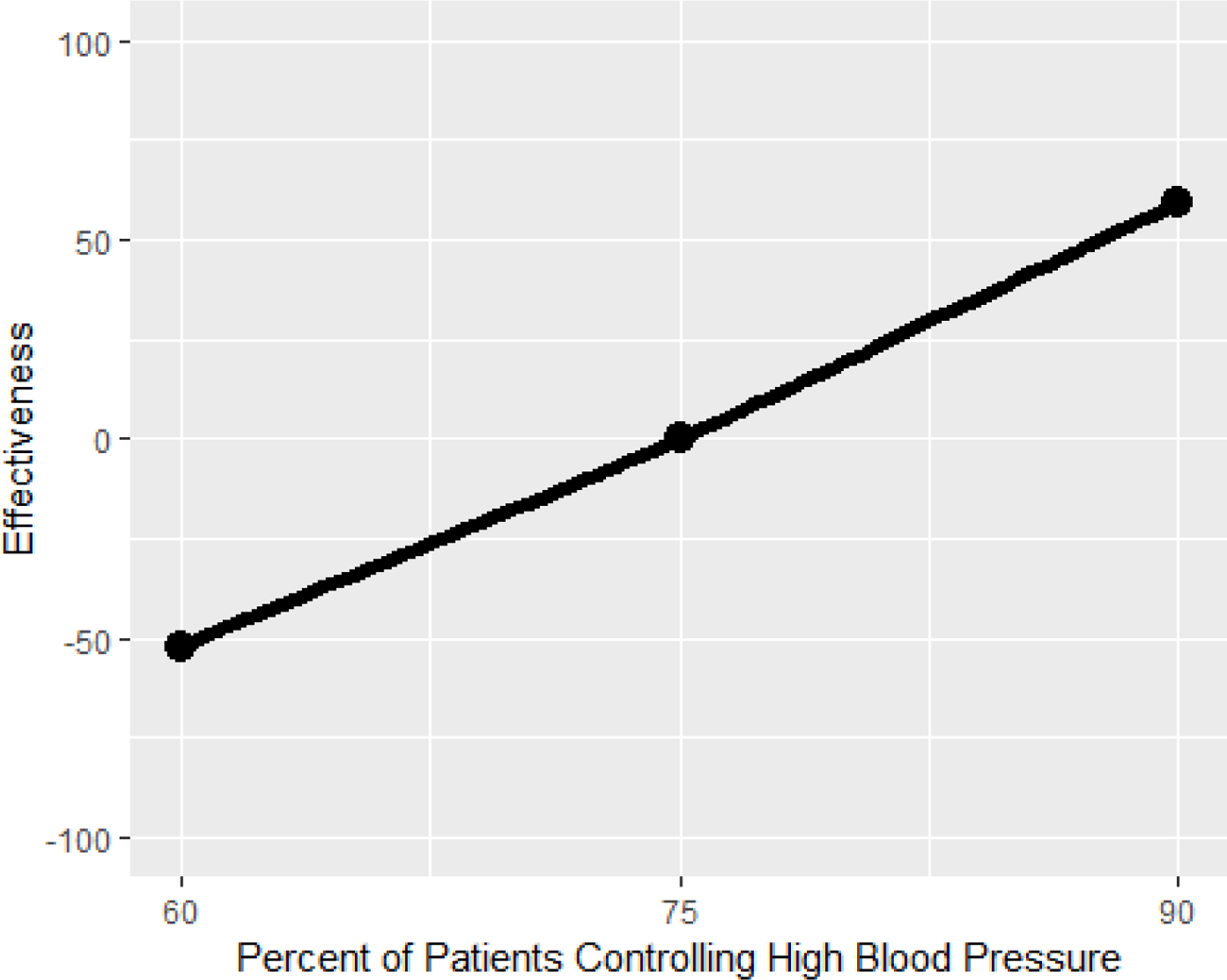

Interpretation. The graph shows that when the percentage of patients controlling their high blood pressure is 75, the effectiveness level is 0; this means the minimum performance is met. If the percentage of patients controlling their high blood pressure decreases to 60, then the effectiveness score decreases to −50. On the other hand, if the rate of controlling high blood pressure increases to 90, the effectiveness score increases to about 60. Data source. eQM

Mnemonic. ihd53h_ec

*Indicator 6. Statin Medication for Patients with Cardiovascular Disease*

Description. This measure is the number of male patients age 21-75 and female patients age 40-75 with cardiovascular disease who had at least one dispensing event for a high or moderate-intensity statin medication (as defined by HEDIS) during the measurement year.

Contingency curve.

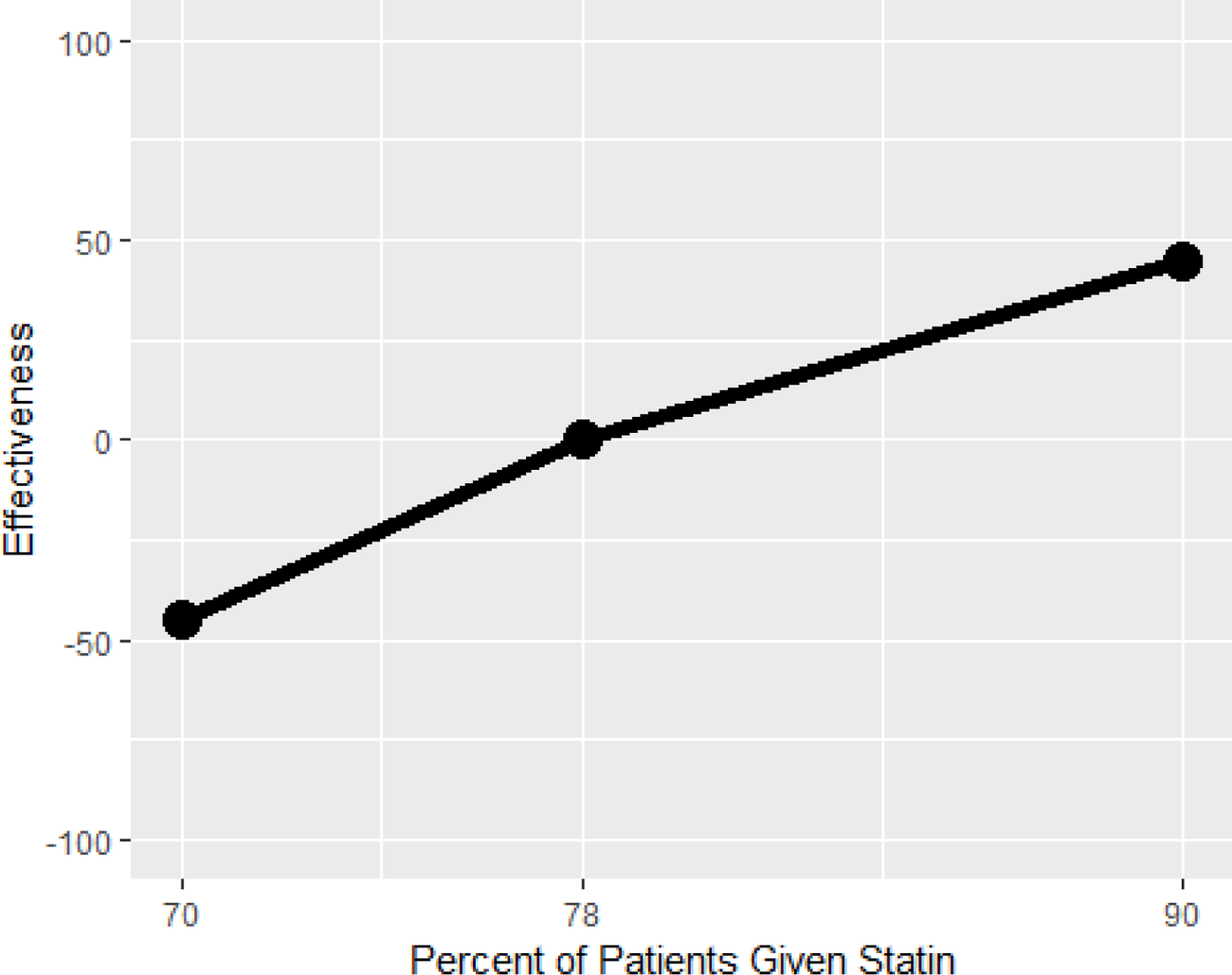

Interpretation. The graph shows that when the percent of patients given a high or moderate-intensity statin medication is 78, the effectiveness level is 0; this means the minimum performance is met. If the percent of patients given statin decreases to 70, then the effectiveness score decreases to about −45. On the other hand, if the percent of patients given statin increases to 90, the effectiveness score increases to about 45.

Data source. eQM

Mnemonic. statn1_ec

*Indicator 7. Effective Continuation Phase Treatment*

Description. This percent is the number of patients over age 18 with a diagnosis of depression who received greater than or equal to 180 days of antidepressant medication through 231 days after the index prescription start date, divided by the number of patients with a diagnosis of depression newly treated with antidepressant medication.

Contingency curve.

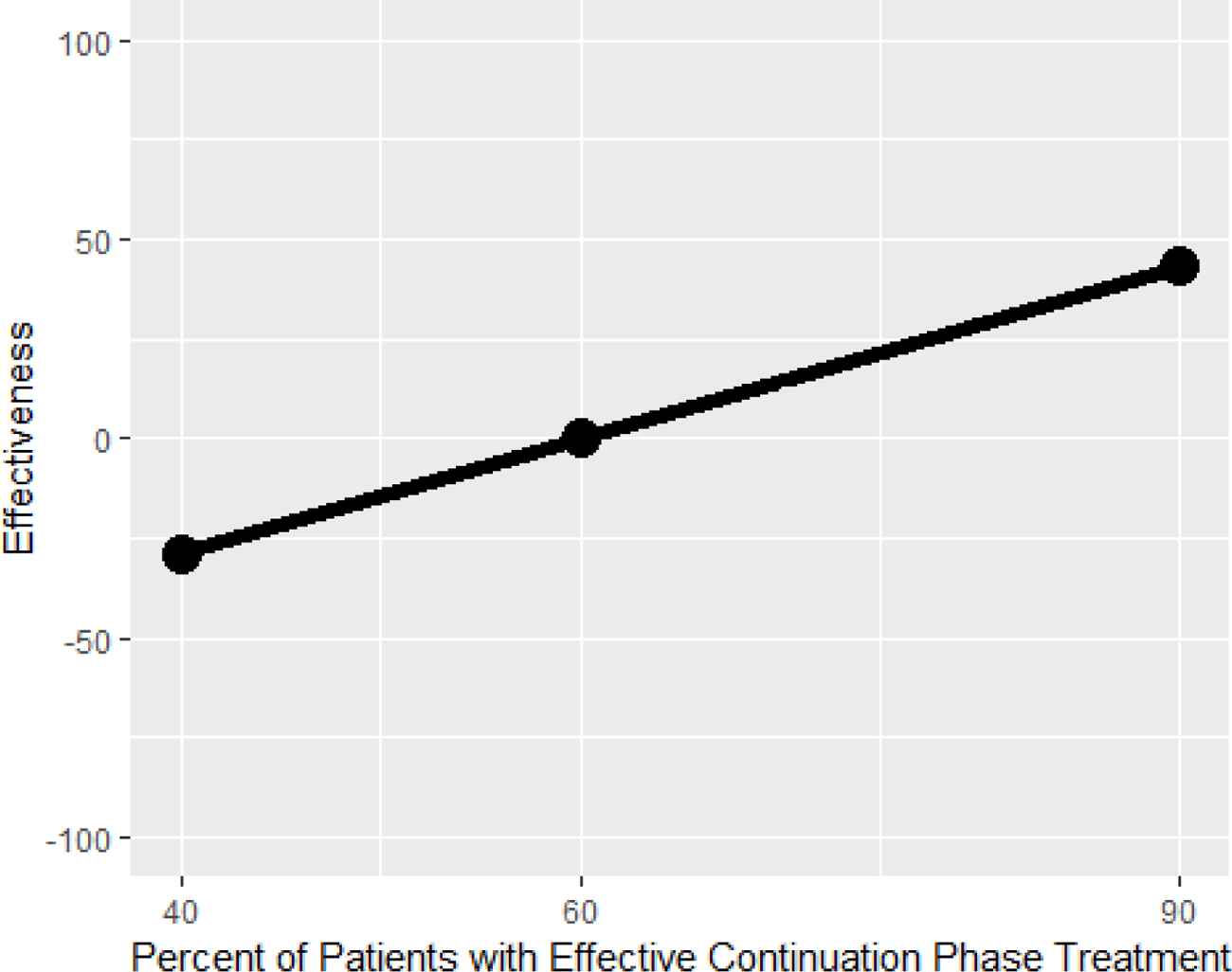

Interpretation. The graph shows that when the percent of patients who received effective continuation phase treatment is 60, the effectiveness level is 0; this means the minimum performance is met. If the percent of patients who received effective continuation phase treatment decreases to 40, then the effectiveness scores decreases to about −20. On the other hand, if the percentage of patients increases to 90, then the effectiveness score increases to about 45.

Data Source. eQM

Mnemonic. mdd47h_ec

*Indicator 8. Renal Testing for Nephropathy*

Description. This measure consists of the percentage of diabetes patients between the ages of 18 and 75 who had a nephropathy screening test during the measurement year.

Contingency curve.

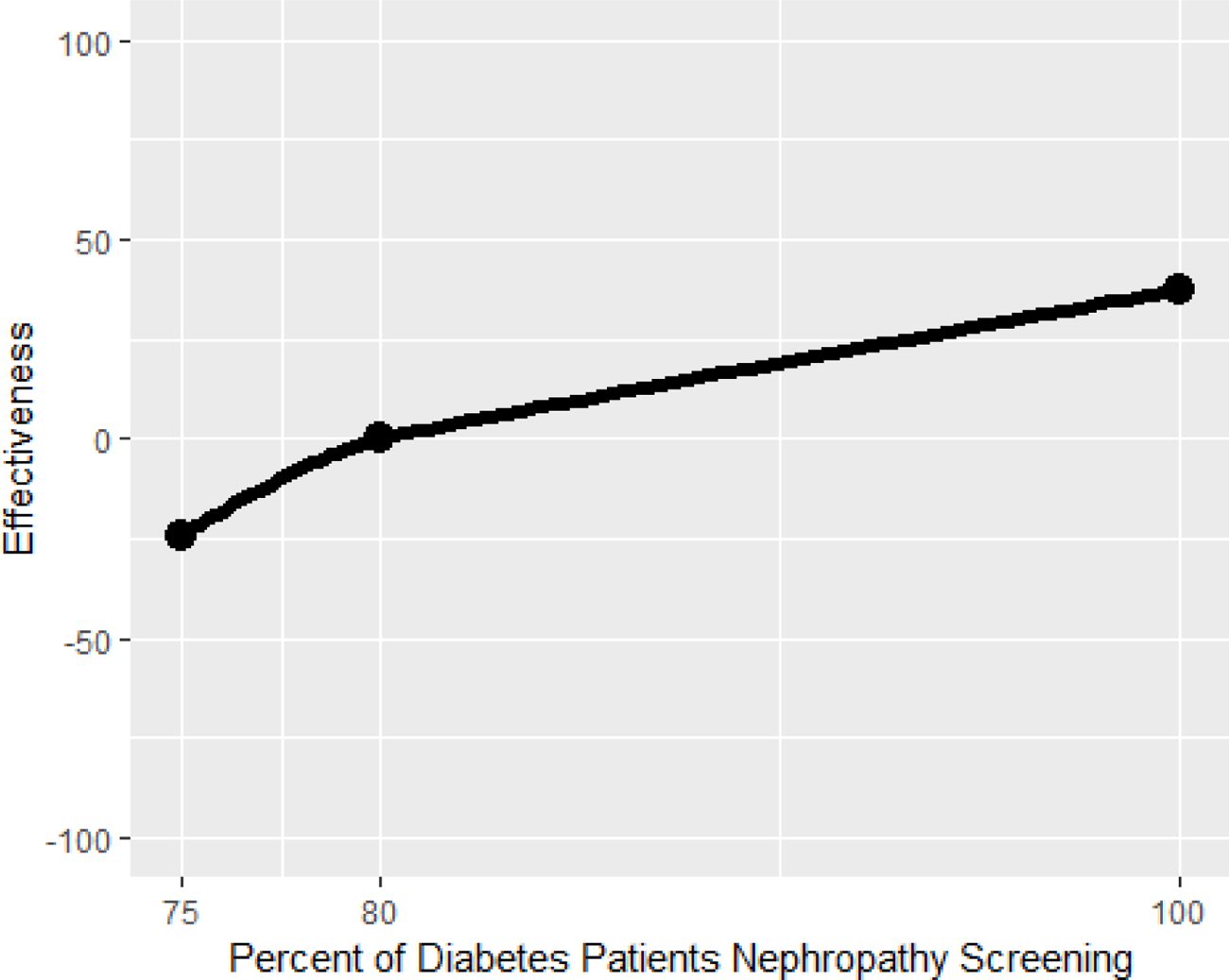

Interpretation. The graph above shows that when the percent of diabetes patients who had nephropathy screening is 80, the effectiveness level is 0; this means the minimum performance is met. If the percent of patients who had nephropathy screening decreases to 75, then the effectiveness score decreases to −25. On the other hand, if the number of nephropathy screening increases to 100, the effectiveness score increases to about 40.

Data source. eQM

Mnemonic. dmg34h_ec

*Indicator 9. Team 2 Day Post Discharge Contact Ratio*

Description. This measure represents the percentage of patients assigned to a given primary care team who were contacted within two days of being discharged (DC) from inpatient care. The post discharge contact is only counted if the individual contacting the patient has a team role of administrative associate, care manager, clinical associate, designated women’s health primary care provider, clinical pharmacist, physician-attending, or primary care provider. Patients are excluded from this measure if they are discharged from an observation specialty and/or are readmitted within two business days to any healthcare facility.

*Note. This measure is also an indicator of trust and also appears under Objective 2*.

Contingency curve.

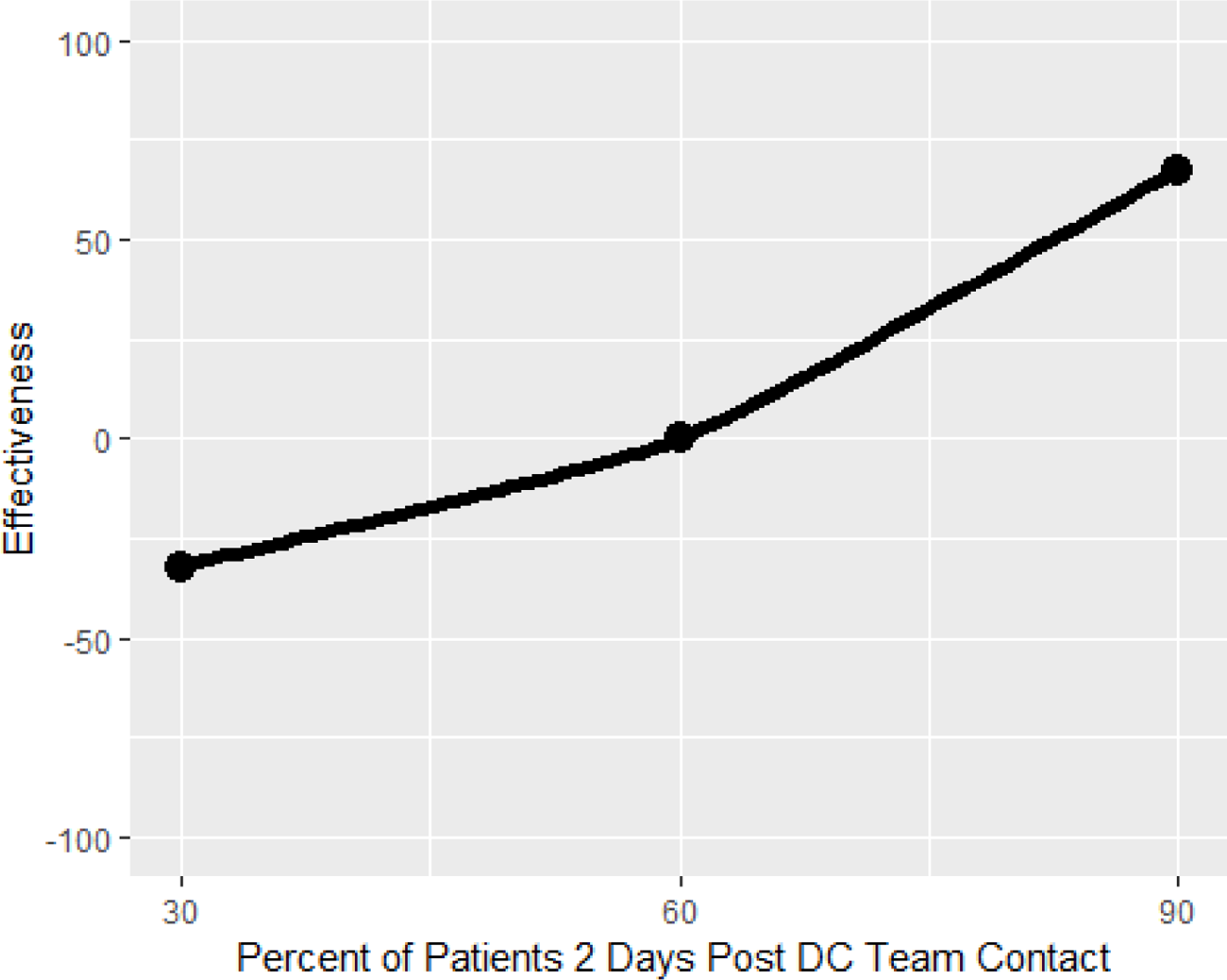

Interpretation. The graph shows that when the percent of patients contacted within two days of being discharged from inpatient care is 60, the effectiveness level is 0; this means the minimum performance is met. If the contact ratio decreases to 30, then the effectiveness score decreases to about −30. On the other hand, if the contact ratio increases to 90, the effectiveness score increases to about 70.

Data source. PACT Compass

Mnemonic. Team 2Day Post DC Contact Ratio – Core Teamlet Only

## Appendix B.2: Suggested measures by objective

Objective 1. Ensure patient has appropriate access to preventive, acute, or chronic health care services when needed.

*Indicator 1.* Average Consult for Community Care

Description. Measure showing average number of referrals and appointments within and outside a specified hospital system to gain access to appropriate specialty service(s).

*Indicator 2.* Timeliness of Community Care Referrals

Description. Timeliness of referrals to specialty care service(s) where an appropriate number of days is assigned to each specialty.

*Indicator 3*. Comprehensive Preventative Visits

Description. Completed preventative care appointments by a patient assigned to a given team during the reporting period. Preventative services could include, but are not limited to, vaccinations, cancer screenings, mammograms, colonoscopies, or stool testing.

*Indicator 4.* Urgent Care Utilization Rate (Adjusted for clinical reason)

Description. Measure should capture why patients utilize urgent care. Utilization rate could be high because patient received ineffective care, do not have access to PCP, or because necessary and reflects good coordination.

*Indicator 1.* Average Effective Partnership Rating

Description. Average rating of providers’ effective partnership. Captured by developing an “Effective Partnership Rating Scale.”

*Indicator 2.* Average Team Trust Rating

Description. Average rating of team trust. Patients could rate how much they trust each member of their assigned primary care team, as well as the overall team. In addition, primary care team members would also rate how much they trust each of their team members, as well as the overall team. This measure is similar to Consumer Assessment of Healthcare Providers and Systems

(CAHPS) Q11, Q14, and Q15 on care coordination and person-centered care but CAHPS does not capture trust.

*Indicator 3.* Effective PC Team Ratio

Description. This measure captures whether a patient’s primary care needs were met by someone from the patients assigned team, when needed. The measure is calculated with the following formula: Number of primary care team encounters WOT (while on team) with patients assigned team member divided by number of primary care team encounter WOT plus the number of ER/Urgent care encounters excluding ED visits in the denominator. This item is similar to PACT 19 in PACT Compass, except PACT 19 includes ED visits.

*Indicator 4.* Continuity Care Ratio

Description. Year over year retention rate with patient panel. Compare across provider, where higher rate means patients are choosing to stay with the provider.

Objective 3. Deliver safe and effective care that comprehensively addresses a given patient’s particular ecological, biological, and/or psychosocial needs.

*Indicator 1*. Consult for Community Care

Description. Percent of referrals to community care that were successfully completed (numerator: number of referrals to community care for which a response from the community care provider was logged into the referring provider’s EHR; denominator: number of referrals to community care logged in the referring provider’s EHR).

*Indicator 2*. Timely Clinic Communication

Description. Mean clinic response time in days to *patient* requests for clinical information and/or the mean clinic response time in days to *provider* requests for clinical information.

*Indicator 3.* Missed Opportunities for Care Coordination

Description. Percent of charts where missed opportunities for care coordination were identified in random peer review process. Could also be measured with number of true trigger positives, e.g., Positive FOBT – no follow up action (colonoscopy) within 60 days, Mammogram with BIRADS 0,4,5 – no follow up action (ultrasound, repeat mammogram, breast biopsy, breast MRI, breast surgery, oncology visit) within 60 days.

*Indicator 4.* Average PCP Safe and Effective Care Rating

Description. This measure captures patients’ average perception of the safe and effective care provided by their primary care provider. Patients rate their primary care provider on a “Safe and Effective Care Scale” which captures patients’ perceptions of whether Objective 3 is being met.

*Indicator 5.* Decrease Inappropriate Antibiotic Prescribing

Description. Number of patients where antibiotics were prescribed for viral URI symptoms divided by number of patients with viral URI symptoms.

## Appendix C: Example of data used to calculate ACSC hospitalization rate per 1,000

| VISN | District | Observed ACSC Admissions | Expected ACSC Admissions | ACSC Patients | Admissions per 1,000 patients | Expected Admissions per 1,000 patients | Observed over Expected Ratio | Risk Standardized Admission Rate per 1,000 (nat avg=24.2) |
| --- | --- | --- | --- | --- | --- | --- | --- | --- |
| 3V10 | District 3 | 6,657 | 7,455 | 307,775 | 21.6 | 24.2 | 0.89 | 21.6 |
| 4V16 | District 4 | 5,355 | 5,870 | 254,960 | 21.0 | 23.0 | 0.91 | 22.0 |
| 1V06 | District 1 | 4,547 | 4,946 | 221,731 | 20.5 | 22.3 | 0.92 | 22.2 |
| 4V19 | District 4 | 3,211 | 3,458 | 156,395 | 20.5 | 22.1 | 0.93 | 22.4 |
| 3V23 | District 3 | 3,217 | 3,456 | 186,317 | 17.3 | 18.5 | 0.93 | 22.5 |
| 5V20 | District 5 | 3,030 | 3,243 | 144,544 | 21.0 | 22.4 | 0.93 | 22.6 |
| 4V17 | District 4 | 4,387 | 4,690 | 222,218 | 19.7 | 21.1 | 0.94 | 22.6 |
| 1V02 | District 1 | 4,191 | 4,311 | 162,583 | 25.8 | 26.5 | 0.97 | 23.5 |
| 2V09 | District 2 | 4,784 | 4,917 | 173,861 | 27.5 | 28.3 | 0.97 | 23.5 |
| 3V15 | District 3 | 4,240 | 4,170 | 152,145 | 27.9 | 27.4 | 1.02 | 24.6 |
| 5V21 | District 5 | 4,720 | 4,602 | 165,259 | 28.6 | 27.8 | 1.03 | 24.8 |
| 1V04 | District 1 | 3,758 | 3,659 | 170,335 | 22.1 | 21.5 | 1.03 | 24.8 |
| 2V07 | District 2 | 5,745 | 5,498 | 260,024 | 22.1 | 21.1 | 1.04 | 25.2 |
| 2V08 | District 2 | 9,579 | 9,163 | 350,783 | 27.3 | 26.1 | 1.05 | 25.3 |
| 5V22 | District 5 | 7,239 | 6,758 | 237,600 | 30.5 | 28.4 | 1.00 | 25.9 |
| 1V01 | District 1 | 3,093 | 2,861 | 137,544 | 22.5 | 20.8 | 1.08 | 26.1 |
| 1V05 | District 1 | 3,807 | 3,469 | 114,204 | 33.3 | 30.4 | 1.10 | 26.5 |
| 3V12 | District 3 | 4,937 | 3,935 | 161,681 | 30.5 | 24.3 | 1.25 | 30.3 |
| VHA Total |  | 86,497 | 86,497 | 3,579,959 | 24.2 | 24.2 | 1.00 | 24.2 |

*Note.* This is an example of the data used by the VHA to calculate ACSC hospitalization rate per 1,000 patients. Specifically, the data includes the observed ACSC admissions, the expected ACSC admissions, and the observed over expected ratio by VA District.

